# Smartphones vs DSLR Cameras in Dental Photography: An In Vitro Assessment of Linear Dimensional Shift in the Esthetic Zone

**DOI:** 10.64898/2026.03.20.26348950

**Authors:** Ankunya Boontharak, Parinya Amornsettachai, Surakit Visuttiwattanakorn

## Abstract

The in vitro study aimed to evaluate linear dimensional shifts in intraoral photographs of the esthetic zone captured using two smartphone cameras—the iPhone 15 Pro Max and the Samsung Galaxy S23 Ultra—compared to a digital single-lens reflex (DSLR) camera, which is regarded as the gold standard for dental photography. Imaging was performed under controlled conditions using a custom-designed stand and stabilizer to maintain a consistent distance and angle between the dental model and the photographic devices. Standardized frontal and occlusal images of the anterior maxillary region were acquired, and point-to-point linear measurements between specified dental landmarks were performed using calibrated digital imaging software.

Each measurement was conducted triple and then averaged across various samples per image to guarantee precision and dependability. Friedman’s test with Bonferroni correction was applied for statistical analysis to evaluate differences among the imaging devices. The results indicated no statistically significant variations in linear measures between the DSLR and the Samsung Galaxy S23 Ultra (p > 0.05), however minor inconsistencies were noted between the DSLR and the iPhone 15 Pro Max. It is important to acknowledge that all images were obtained utilizing the stabilization system, which contrasts with the conventional handheld approach applied in clinical environments and could impact the external validity of the results. The Samsung Galaxy S23 Ultra, in telephoto mode, demonstrated measurement precision similar to that of a DSLR camera, potentially serving as a reliable choice for clinical intraoral photography. The iPhone 15 Pro Max demonstrated potential, although minor measurement discrepancies.

## 1. Introduction

### Background and Rationale

Aesthetics are an important aspect of nowadays dental practice. To effectively address the patient’s primary aesthetic concerns, each treatment procedure requires a well-organized plan, appropriate technique, and clear communication. Accurate measurement of tooth proportions is especially important in the anterior region of cosmetic dental restoration. Efficient communication among the patient, practitioner, and dental technician is essential for achieving aesthetic goals. In this context, photography serves a crucial function, particularly for documentation purposes [1–5]. This became increasingly apparent during the COVID-19 epidemic, when internet communication replaced face-to-face encounters. With the development of virtual medical consultations, digital photography emerged as an essential instrument for remote diagnosis, education, and patient communication [6, 7].

In recent years, digital technology has progressively influenced all aspects of dentistry. Digital cameras and dental photography have become essential instruments, especially in aesthetic dentistry. Nonetheless, their applications encompass not just cosmetics but also specializations such as orthodontics, periodontics, implantology, dental technology, and oral surgery [8]. Photographic data serves as a “objective and efficient communication tool among dentists, patients, and technicians,” vital for treatment planning, smile design, and mock-up processes [9, 10]. Traditional DSLR cameras fitted with a macro lens (e.g., 105 mm) are regarded as the gold standard for obtaining 1:1 (life-size) proportion measurements in dentistry photography [11].

Nevertheless, recent smartphone cameras have advanced rapidly, with numerous models providing image quality comparable to that of professional cameras. The use of smartphone cameras in dental photography has only recently begun to appear in the literature [12–22]. Their lightweight design, intuitive user interface, and accessibility make smartphones a potentially valuable tool in dental practice [23].

Therefore, the purpose of this study is to evaluate and compare the linear dimensional shifts in dental image proportions captured using a DSLR camera versus those taken with advanced smartphone cameras.

## 2. Reviews of related literatures

### Image Distortion

Geometric distortion (GD) in digital still cameras (DSCs) mostly results from inconsistencies in magnification across the image field generated by the camera lens. The most obvious demonstration of this distortion is the modification of straight lines to provide an appearance of curvature. Consequently, the proportional relationships with the parts of the image are altered, posing significant challenges in applications involving natural environments, architectural photography, or portraits. Distortion is mathematically characterized by a two-dimensional (2D) displacement map, which delineates the divergence of each point in a distorted image from its equivalent position in an ideal, undistorted image. The image’s center is generally regarded as distortion-free, with the magnification at this location determining the camera’s effective focal length. Geometric distortions are categorized according to the variation in magnification radially from the center of the image. The predominant types consist of barrel distortion, characterized by a reduction in magnification as one proceeds out from the center, and pincushion distortion, where magnification increases radially. Distortion forms that do not adhere strictly to these definitions are known as wave distortion [24].

Positive radial displacement, or barrel distortion, is a geometric aberration found in wide-angle and zoom lenses. Spherizing occurs when straight lines, especially near the image’s boundaries, appear bent outward. Image may appear projected onto a convex surface. Architectural or grid-like photos show this effect, when straight lines curve, narrowing the top and bottom and expanding the center. Barrel distortion (Fig 1)is caused by decreasing magnification as objects travel away from the lens’ optical axis. Shorter focal lengths increase distortion. Barrel distortion is a major problem in dental photography, particularly when the camera is placed too close to the subject. The accuracy of clinical documentation may be jeopardized due to altered anatomical proportions caused by this closeness. Increasing the distance between the subject and the camera is crucial to counteracting this effect. Therefore, taking precise, distortion-free pictures in a dental office requires careful handling and positioning of both DSLR and smartphone cameras.

**Fig 1.**
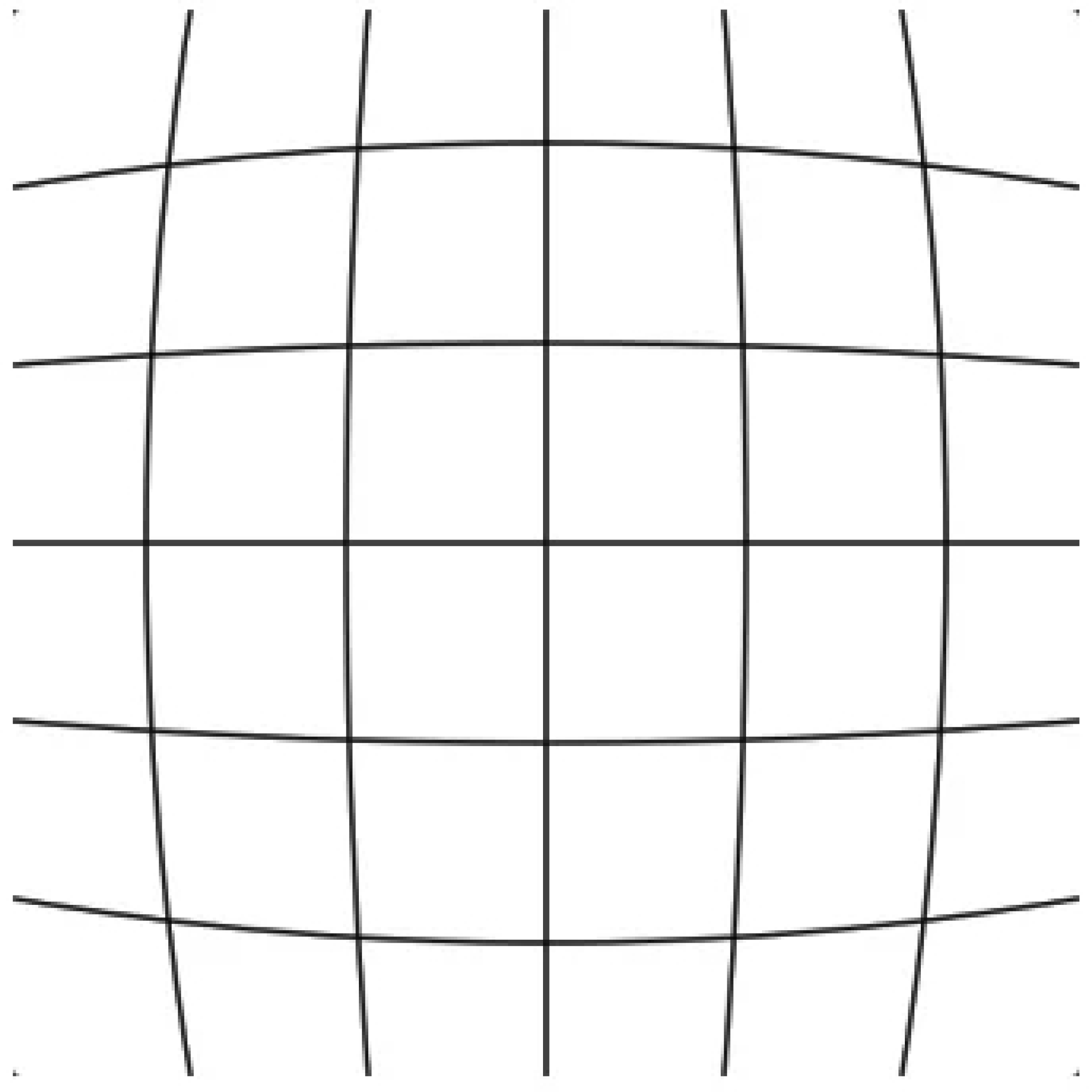
Barrel Distortion.

The geometric opposite of barrel distortion is pincushion distortion, also known as negative radial displacement (Fig 2). This type of distortion bends lines inward toward the image’s center rather than outward. This type of distortion is particularly common when photographing with telephoto lenses or at the telephoto end of a zoom range, with focal lengths ranging from 85 mm to 105 mm. Subjects appear compressed and obviously pinched as a result of this distortion, which worsens as magnification increases near the image’s boundaries.

**Fig 2.**
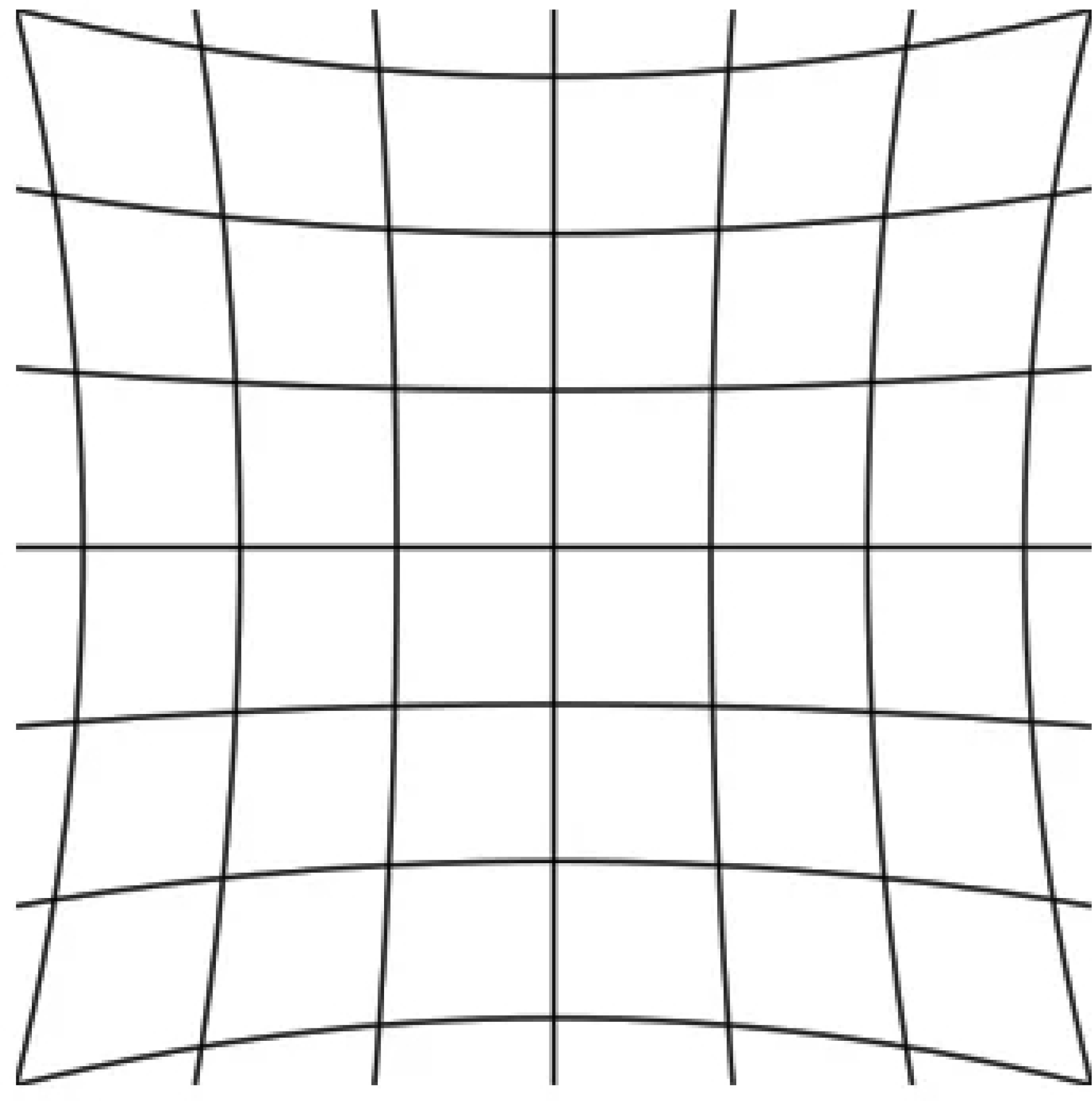
Pincushion distortion.

Mustache distortion, also known as wave distortion (Fig 3), combines the characteristics of both barrel and pincushion distortion. Despite being uncommon, mustache distortion is particularly challenging to solve due to its complicated nonlinear structure.

**Fig 3.**
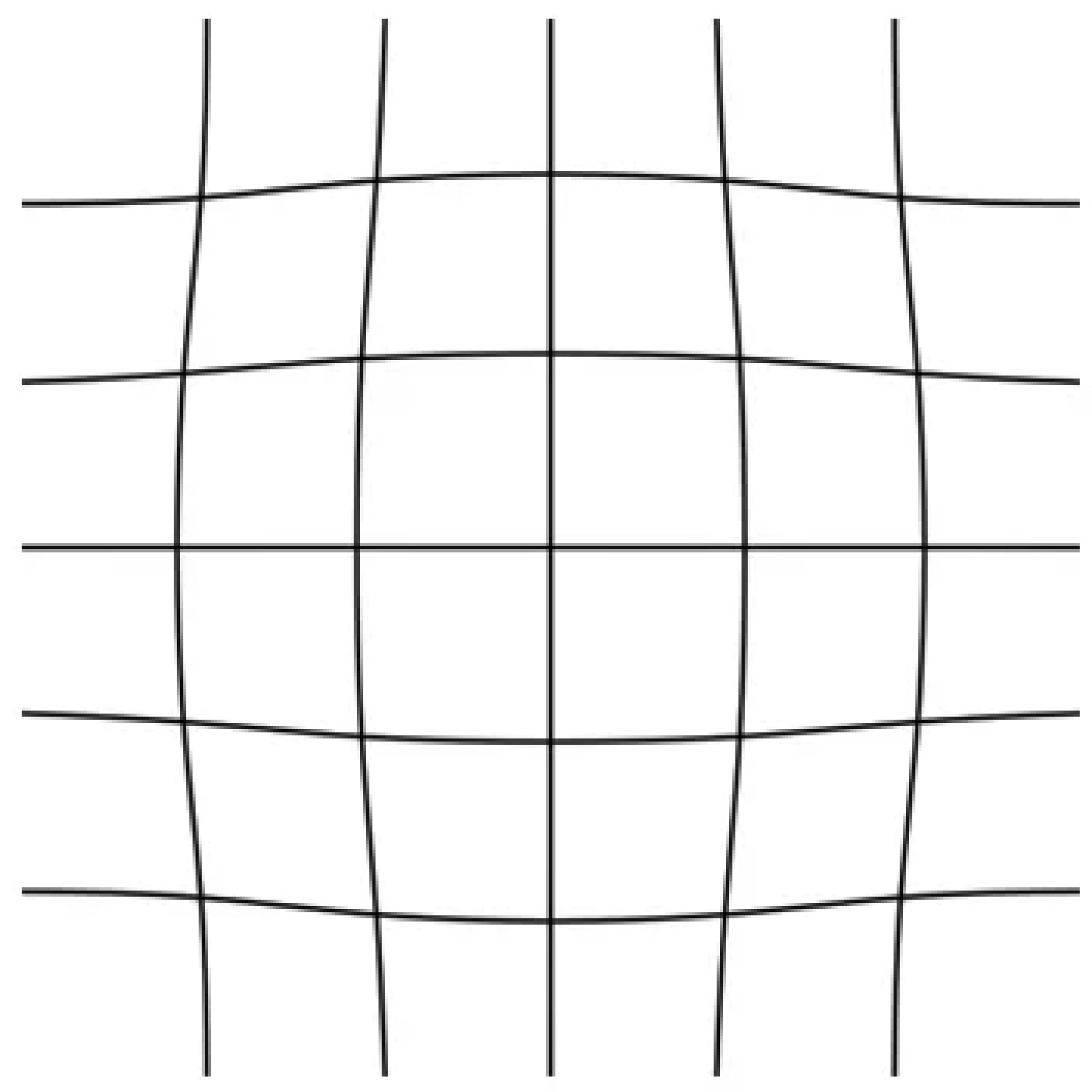
Mustache distortion.

### Instrument for tooth proportion measurement

#### Digital cameras and imaging systems

Photography is based on the relationship between light and a photosensitive substance. The creation of photographic results, especially in therapeutic contexts, requires precise regulation and manipulation of light, and also systematic control of key variables. These include the technological features of the camera equipment, image composition, exposure parameters, sharpness, and accurate color balance. In both general and specialized contexts, these elements are crucial for ensuring the accuracy and diagnostic value of photographic documentation.

#### Digital camera

A digital single-lens reflex (DSLR) camera combines a single-lens reflex system with a digital image sensor. In this configuration, light traverses the camera lens and is reflected by a mirror, channeling the picture either to a prism or to the image sensor. The image is observed directly through the lens using the optical viewfinder when directed to the prism. In contrast, when the sensor captures the image, it is presented on a digital display. In both instances, the optical viewfinder and digital display convey fundamentally identical images.

Digital cameras are typically categorized into three classifications based on their attributes and functionalities: amateur, semi-professional, and professional. Professional DSLR systems are regarded as the benchmark for generating high-quality, consistent intraoral images [1, 25].

They provide considerable flexibility to modify image settings based on clinical needs and environmental factors. These systems exhibits exceptional compatibility with macro lenses, facilitating high-magnification close-up imagery, constant white balance, and tailored flash systems for dental macro-photography [26]. Semi-professional DSLR cameras exhibit comparable designs but possess worse viewfinders, restricted lens interchangeability, and streamlined controls, rendering them less suitable for specialized clinical photography.

Despite their benefits, DSLRs may exhibit limitations in dental photography, such as potential image distortion, shot inconsistency, depth-of-field issues, inadequate flash positioning, misalignment of the focusing plane, extended focusing durations, and fixed exposure or flash settings in specific configurations [1].

In recent years, cameras have become more integrated into mobile devices, with smartphone cameras currently being the most prevalent imaging systems worldwide. Technological progress in this sector has surged swiftly, propelled by a competitive marketplace and substantial customer demand. The primary advantages of smartphone cameras encompass their compact size, lightweight construction, user-friendliness, integrated image processing, various computational imaging techniques, wireless connectivity, and the ability to generate high-resolution images [27].

Nevertheless, smartphone cameras frequently lack extensive manual capabilities. Many models depend significantly on automatic modifications according to ambient lighting, which may result in fluctuations in light exposure if the sensor identifies inconsistent lighting conditions [27]. In response, certain manufacturers have lately implemented AE/AF lock functions, facilitating more control and consistency in digital photography.

#### Lens selection

In dental photography, the macro lens is predominantly employed for its capacity to generate high-resolution close-up images vital for clinical documentation. These lenses provide various beneficial features, such as image stabilization, fixed focal lengths, enhanced image quality, and rapid autofocus capabilities [26]. A macro lens with a focal length ranging from 80 mm to 105 mm is especially efficient, enabling 1:1 magnification while reducing image distortion [10, 25, 26]. The prevailing method for obtaining clinical dental photographs is the utilization of a DSLR camera paired with a 100 mm macro lens and a macro ring flash [28, 29]. This combination facilitates precise full-arch imaging by maintaining proportional relationships in the image and ensuring minimal distortion [26].

#### Flash

Flash is an important part of dental photography, as it ensures proper light balance for shutter speed, aperture, and ISO settings. A ring flash is commonly used to deliver optimal uniform light and reduces shadows for intraoral photography when using a smaller aperture (higher f-stop), as an aperture of f/22 is frequently recommended. While f/10 is frequently recommended for extraoral photography to increase light intake while preserving suitable image clarity [30].

#### File

A computer-based representation of visual data, a digital image has benefits including instant viewing and electronic transmission. An electronic sensor, such as a complementary metal-oxide-semiconductor (CMOS) sensor or a charge-coupled device (CCD), is usually used to capture light to create digital photographs [31]. Compared to CMOS sensors, CCD sensors typically offer higher sensitivity and better image quality; however, they are expensive and require more power to function. Pixel arrangements in a matrix make up the graphical composition of digital image files [31]. Overall imaging performance has significantly improved as smartphone camera technology has advanced from early low-resolution CCD sensors to contemporary high-resolution CMOS sensors [32]. There are several formats in which digital photos can be stored. Vector-based formats used in illustration software and bitmap-based formats such as JPEG, HEIC, GIF, TIFF, and BMP are common examples [31, 33]. Due to its ability to allow data interchange between systems and support picture modification, the Digital Imaging and Communications in Medicine (DICOM) standard is widely used in medical imaging [31].

## 3. Objectives

### Research Question

Is there any difference in dental image proportions of linear dimensional shift using different smartphone cameras compared with a DSLR camera?

### Objectives

To measure and compare the dental image proportions of linear dimensional shift when using different smartphone cameras compared with a DSLR camera?

### Research Hypothesis

Different smart devices, including DSLR and smartphone cameras, affect the linear dimensional shift in dental image proportions.

### Statistical Hypothesis

To compare the linear dimensional shift in dental image proportions when using smartphone cameras compared with a DSLR camera.

H0: There is no difference in the linear dimensional shift in dental image proportions when using different smartphone cameras compared with a DSLR camera.

H1: There is a difference in the linear dimensional shift in dental image proportions when using different smartphone cameras compared with a DSLR camera.

## 4. Research design and methodology

### Study Design

In Vitro Experimental Study

### Conceptual Framework

**Fig 4.**
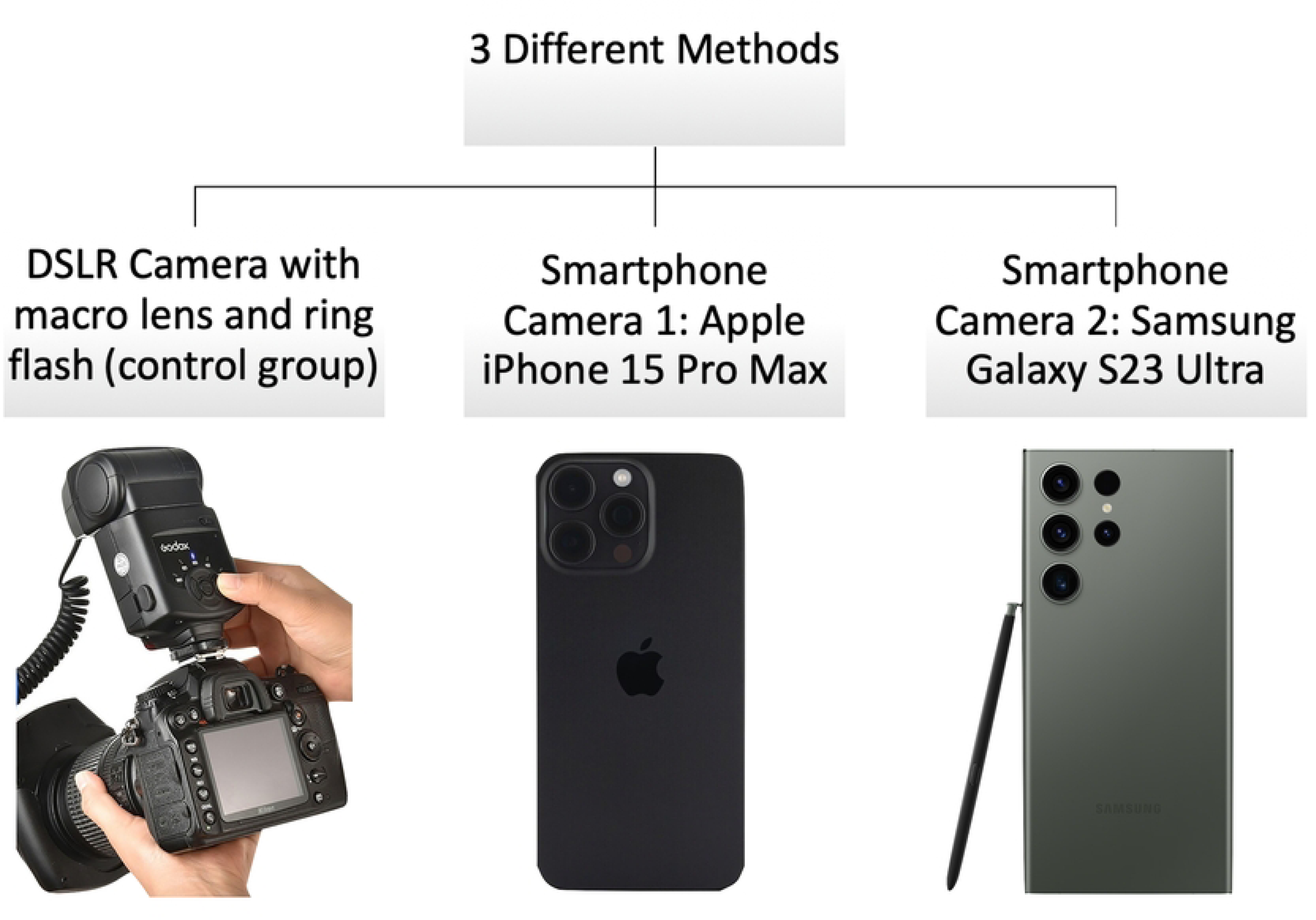
Three different methods on this study.

**Table 1.** Further information of each method used in this study.

| No | Method | Description |
| --- | --- | --- |
| 1 | DSLR camera with macro lens and ring flash (control group) | DSLR camera: Nikon D800 with macro lens 105 mm F2.8, Nikon®, Japan<br>Ring flash: Godox®, China |
| 2 | Smartphone 1 | Smartphone camera: iPhone 15 Pro Max, Apple® USA |
| 3 | Smartphone 2 | Smartphone camera: Samsung Galaxy S23 Ultra, Samsung®, South Korea |

### Instrument and software used in the study

**Table 2.**
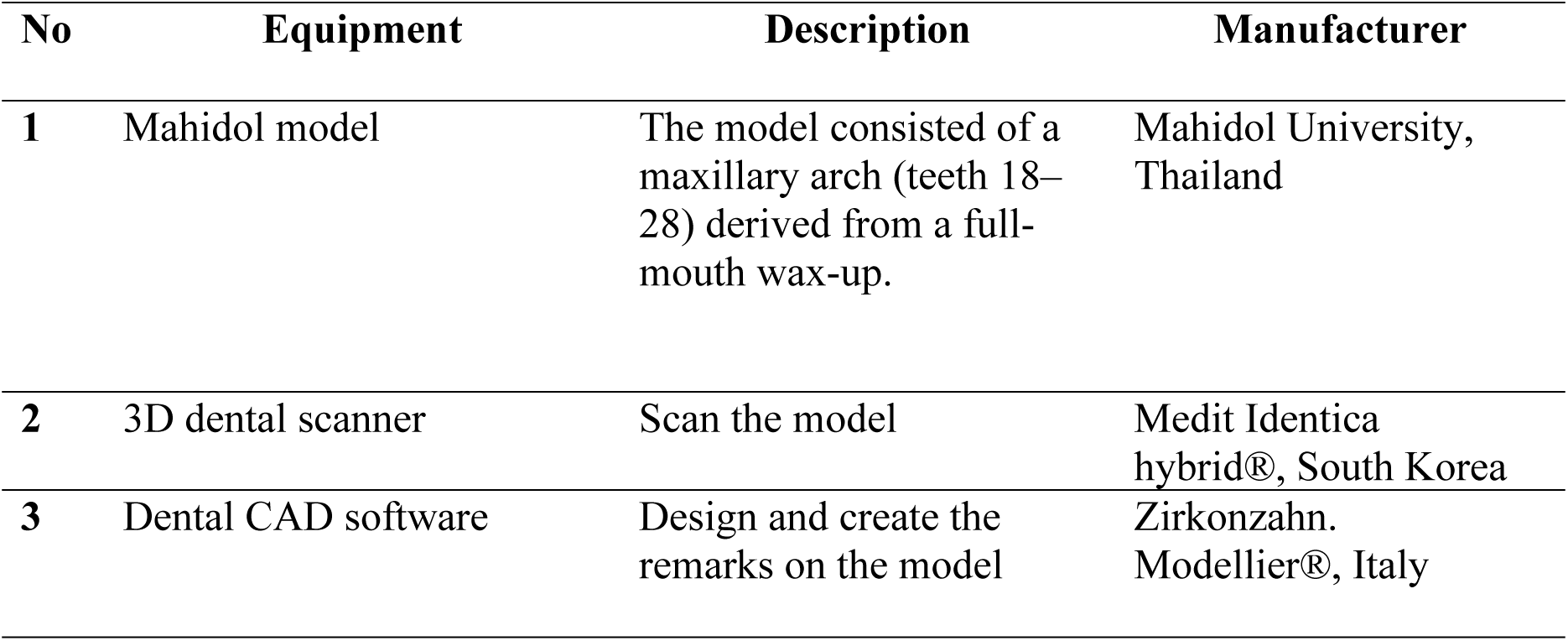
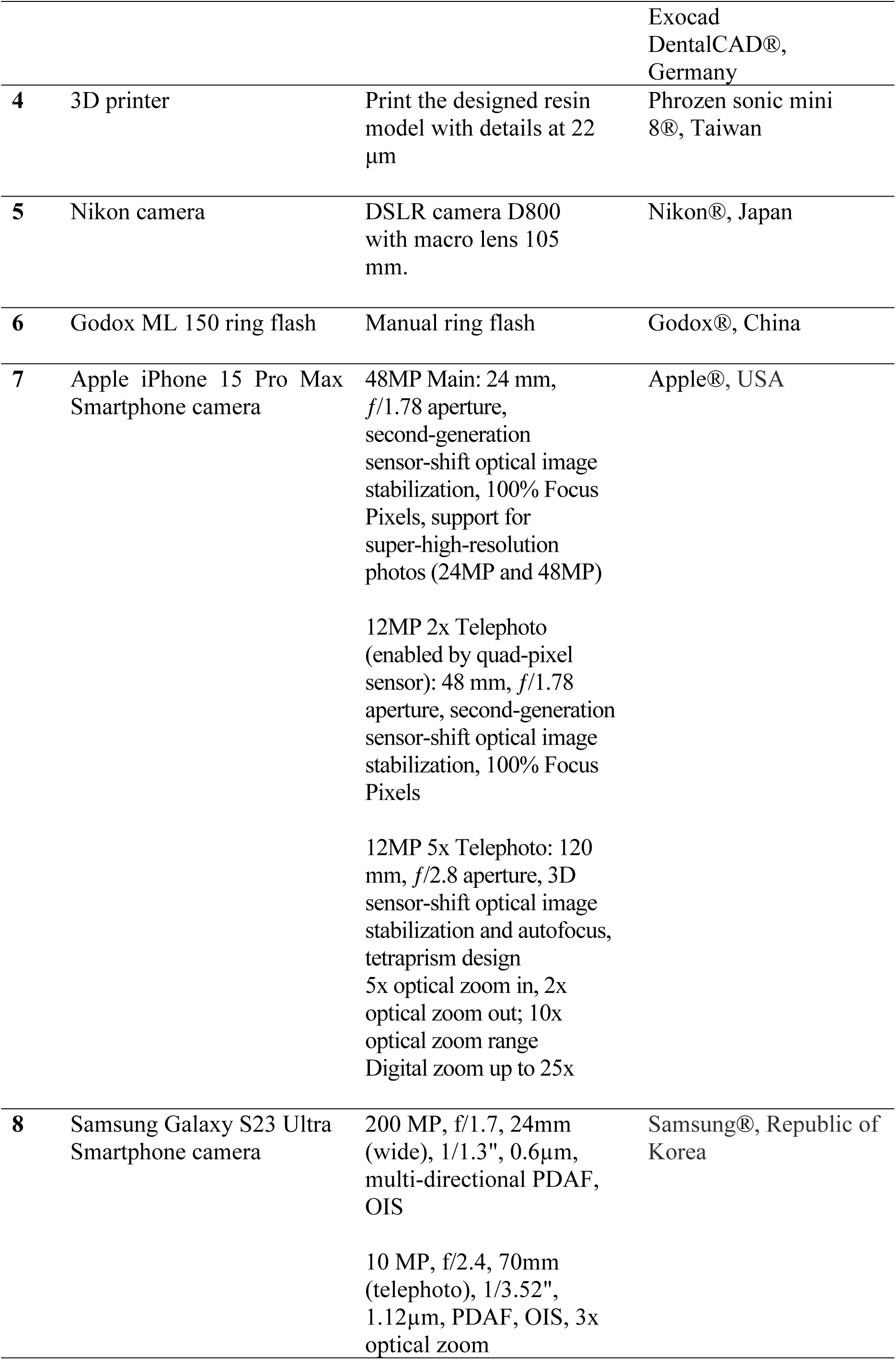
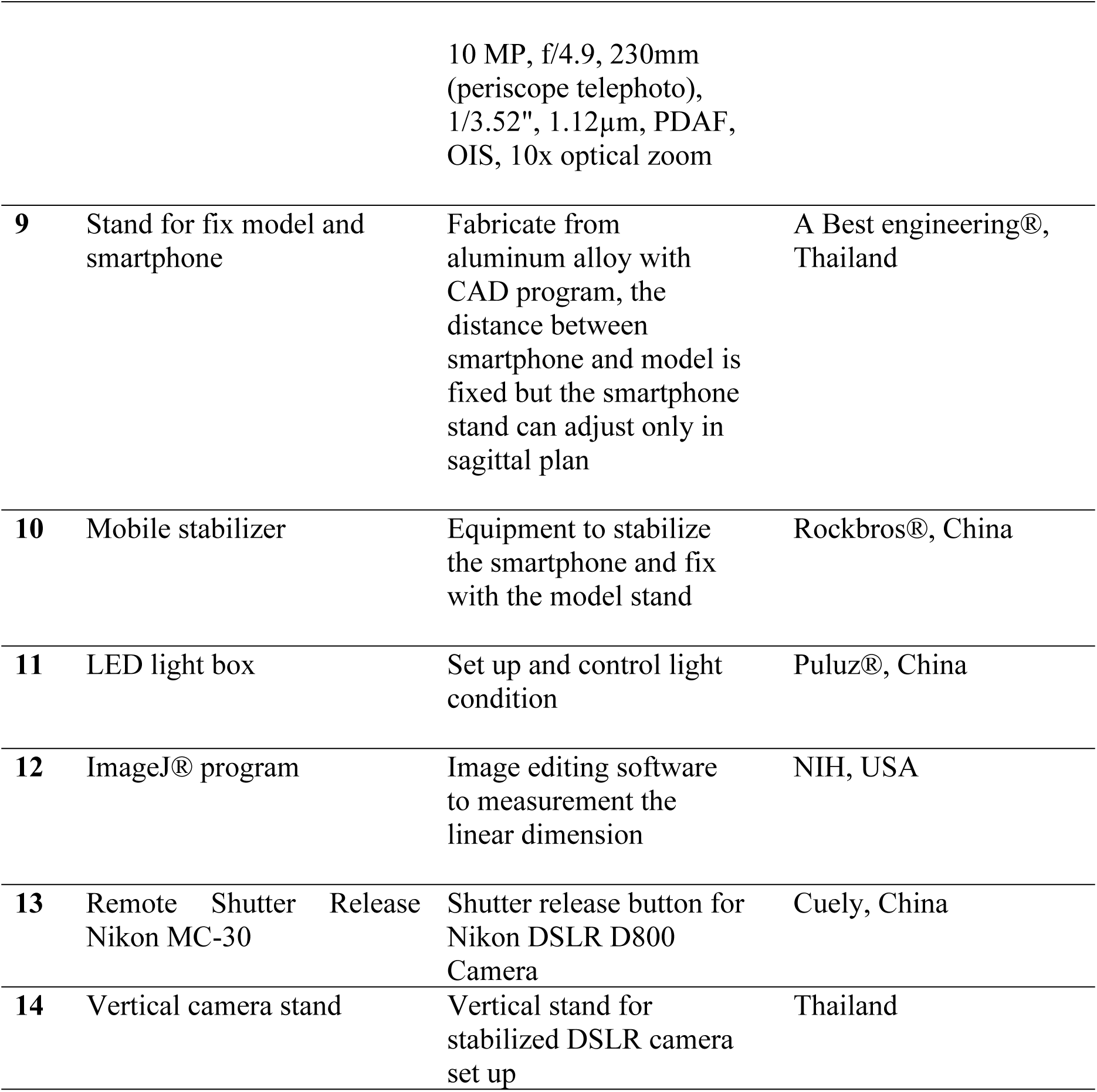
The equipment and software used in the study.

| No | Equipment | Description | Manufacturer |
| --- | --- | --- | --- |
| 1 | Mahidol model | The model consisted of a maxillary arch (teeth 18–28) derived from a full-mouth wax-up. | Mahidol University, Thailand |
| 2 | 3D dental scanner | Scan the model | Medit Identica hybrid®, South Korea |
| 3 | Dental CAD software | Design and create the remarks on the model | Zirkonzahn. Modellier®, Italy |
|  |  |  | Exocad<br>DentalCAD®,<br>Germany |
| 4 | 3D printer | Print the designed resin model with details at 22 $\mu\text{m}$ | Phrozen sonic mini 8®, Taiwan |
| 5 | Nikon camera | DSLR camera D800 with macro lens 105 mm. | Nikon®, Japan |
| 6 | Godox ML 150 ring flash | Manual ring flash | Godox®, China |
| 7 | Apple iPhone 15 Pro Max Smartphone camera | 48MP Main: 24 mm, $f/1.78$ aperture, second-generation sensor-shift optical image stabilization, 100% Focus Pixels, support for super-high-resolution photos (24MP and 48MP)<br>12MP 2x Telephoto (enabled by quad-pixel sensor): 48 mm, $f/1.78$ aperture, second-generation sensor-shift optical image stabilization, 100% Focus Pixels<br>12MP 5x Telephoto: 120 mm, $f/2.8$ aperture, 3D sensor-shift optical image stabilization and autofocus, tetraprism design<br>5x optical zoom in, 2x optical zoom out; 10x optical zoom range<br>Digital zoom up to 25x | Apple®, USA |
| 8 | Samsung Galaxy S23 Ultra Smartphone camera | 200 MP, $f/1.7$, 24mm (wide), $1/1.3''$, $0.6\mu\text{m}$, multi-directional PDAF, OIS<br>10 MP, $f/2.4$, 70mm (telephoto), $1/3.52''$, $1.12\mu\text{m}$, PDAF, OIS, 3x optical zoom | Samsung®, Republic of Korea |
|  |  | 10 MP, f/4.9, 230mm (periscope telephoto), 1/3.52", 1.12µm, PDAF, OIS, 10x optical zoom |  |
| 9 | Stand for fix model and smartphone | Fabricate from aluminum alloy with CAD program, the distance between smartphone and model is fixed but the smartphone stand can adjust only in sagittal plan | A Best engineering®, Thailand |
| 10 | Mobile stabilizer | Equipment to stabilize the smartphone and fix with the model stand | Rockbros®, China |
| 11 | LED light box | Set up and control light condition | Puluz®, China |
| 12 | ImageJ® program | Image editing software to measurement the linear dimension | NIH, USA |
| 13 | Remote Shutter Release Nikon MC-30 | Shutter release button for Nikon DSLR D800 Camera | Cuely, China |
| 14 | Vertical camera stand | Vertical stand for stabilized DSLR camera set up | Thailand |

### Model preparation

A maxillary arch model including teeth numbers 18 to 28, derived from a full-mouth wax-up provided by Mahidol University, was scanned using a 3D scanner (Medit Identica Hybrid®, Republic of Korea). The scanned data were then imported into CAD software (Zirkonzahn.Modellier®, Italy), where the model was positioned within the program’s digital articulator, as illustrated in Fig 5. The occlusal plane was aligned using the smallest grid line reference view.

**Fig 5.**
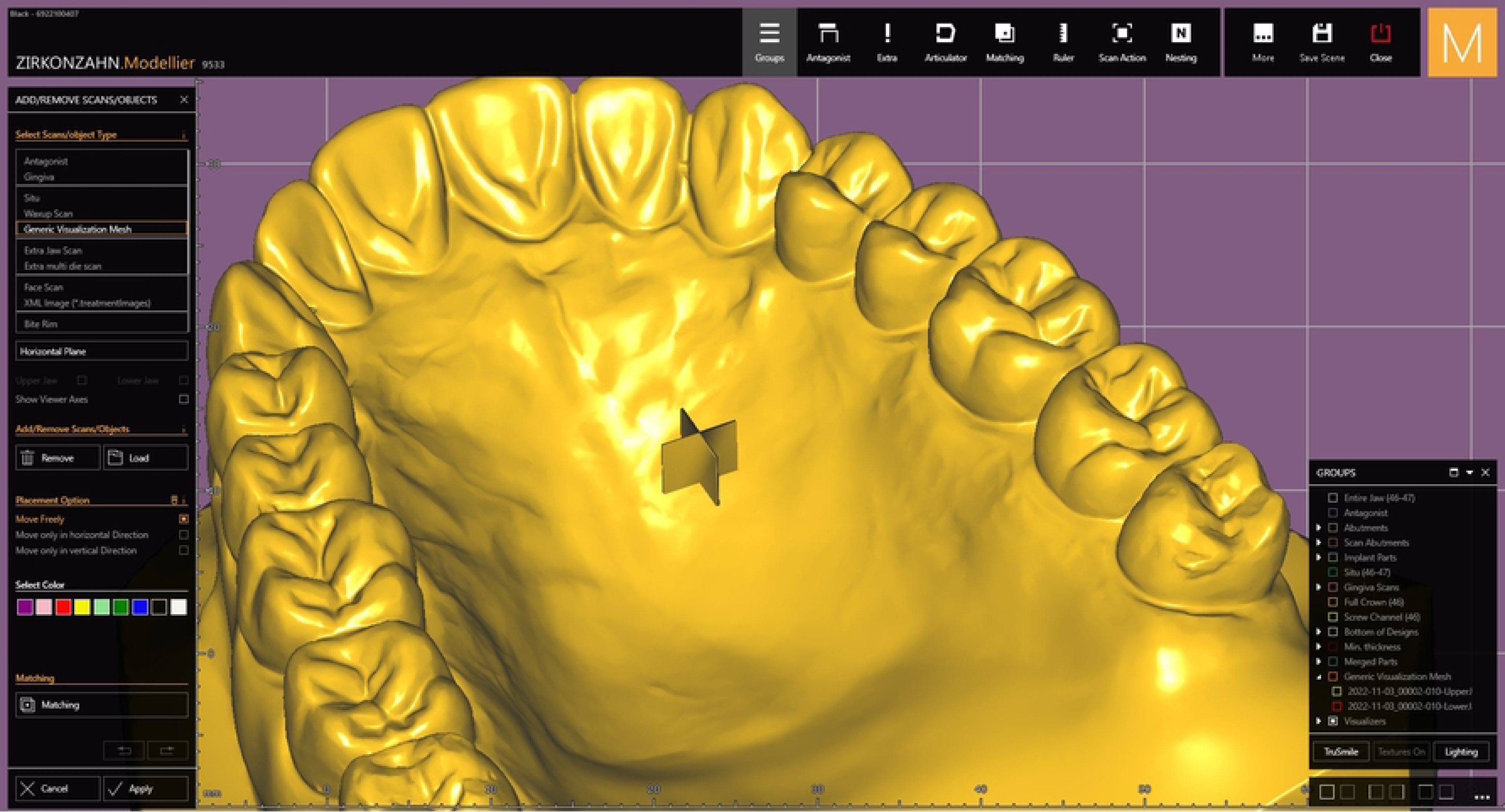
Scanned model in CAD software program.

A reference marker in the shape of a plus sign (“+”) measuring 5 × 5 mm with an approximate line thickness of 0.3 mm was created and digitally merged onto the model at the intersection of the grid lines along the midline, positioned between the upper right and upper left central incisors. This marker served as a reference point in both the frontal and occlusal views. The center of a photograph is generally considered to exhibit minimal distortion compared to its peripheral areas. In this study, the centrally positioned 5 × 5 mm “+” marker, located at the midline between the upper right central incisor and the upper left central incisor, served as the reference point for all photographic images. The central plus mark in the frontal view was labeled as “Control 1” and in the occlusal view as “Control 2.” These markers were used to calibrate pixel dimensions and linear measurements across images obtained from different devices. To ensure consistency, the central “+” marker was aligned with the center of the photographic frame in all images.

Smaller replicas of the original marker were subsequently duplicated, scaled down, and positioned on the labial surfaces from the upper right canine to the upper left canines. These were placed at the incisal third and gingival third regions, aligned with the grid line intersections. The placement of these markers provided consistent reference points in both frontal and occlusal views, as demonstrated in Figs 6 and 7.

**Fig 6.**
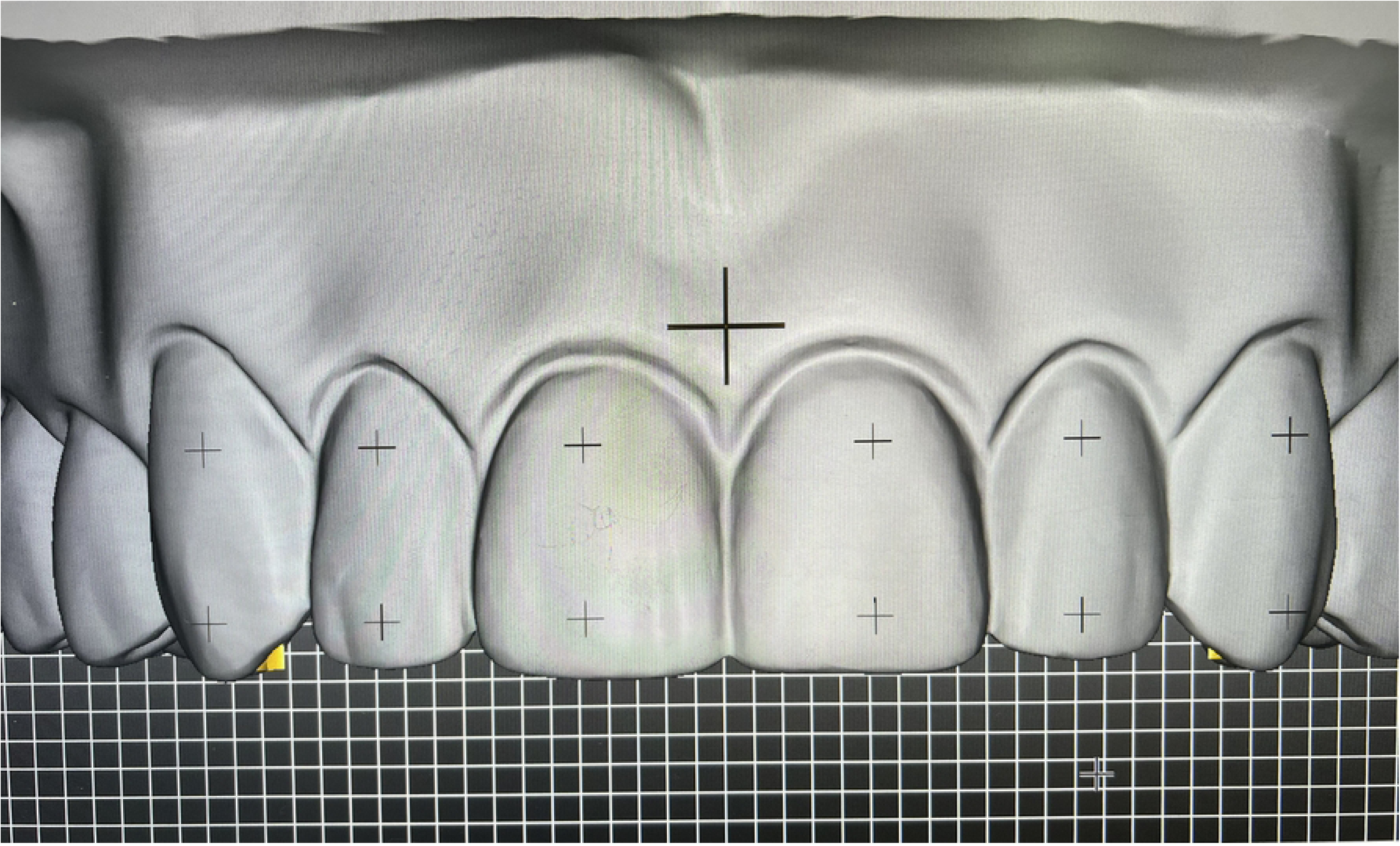
The plus marks were merged on the teeth surface on the frontal view.

**Fig 7.**
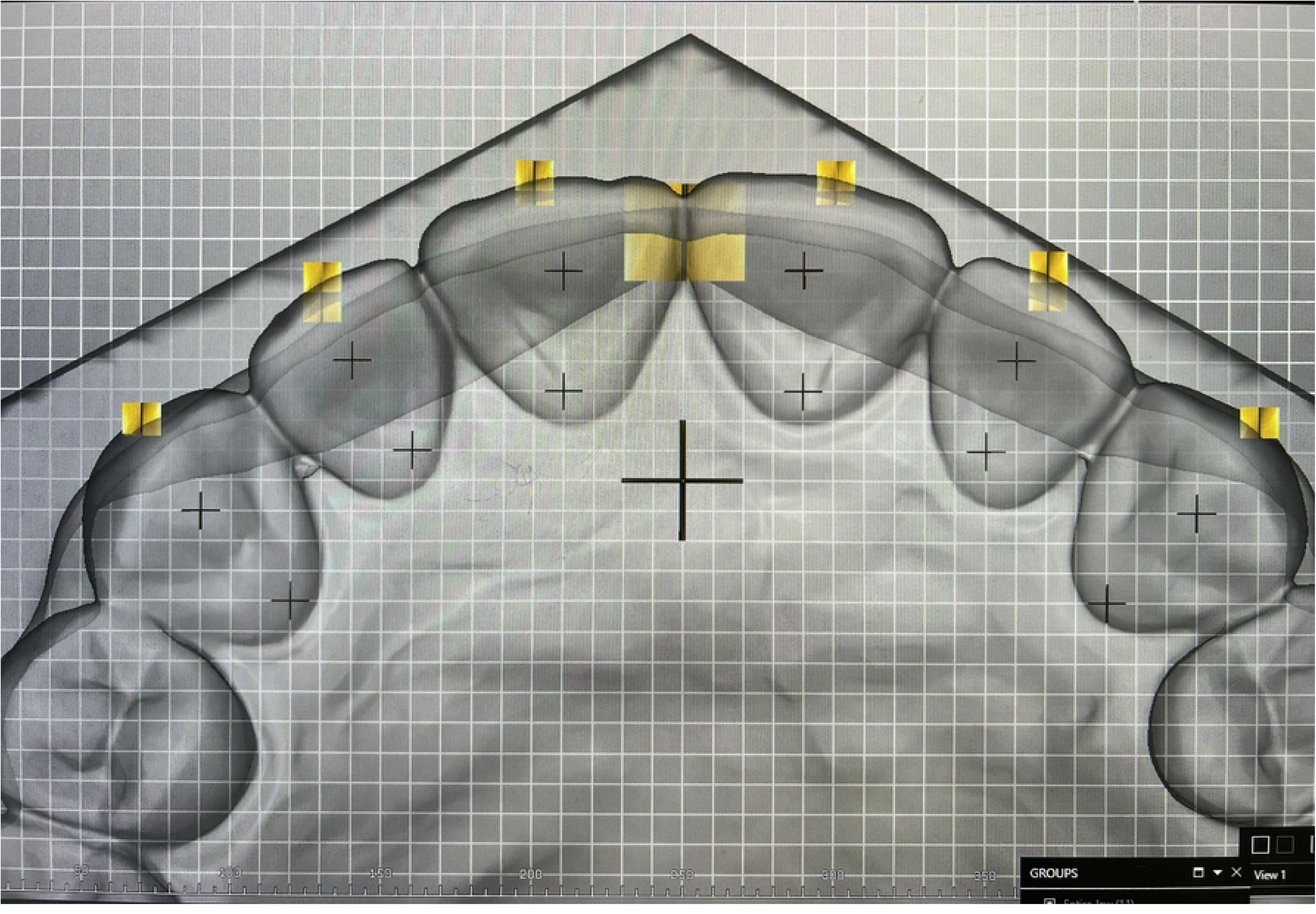
The plus marks were merged on the teeth surface on the occlusal view.

The model with integrated reference markers was fabricated using a 3D printer (Phrozen Sonic Mini 8K®, Taiwan) and gray resin. An 18% gray resin was specifically selected to minimize light scattering during subsequent optical analysis. On the labial surfaces of teeth 13, 12, 11, 21, 22, and 23, additional “+” markers measuring 1 × 1 mm with an approximate line thickness of 0.1 mm were affixed at the center of both the cervical third and incisal third regions. In the frontal view, these markers were designated as reference points A through L, as illustrated in Fig 8. Additionally, “+” markers were placed on the palatal surfaces of the upper anterior teeth, extending from the right canine to the left canine. These markers were positioned at the center of the cervical third and incisal third regions. In the occlusal view, these points were labeled as reference points M through X, as shown in Fig 9.

**Fig 8.**
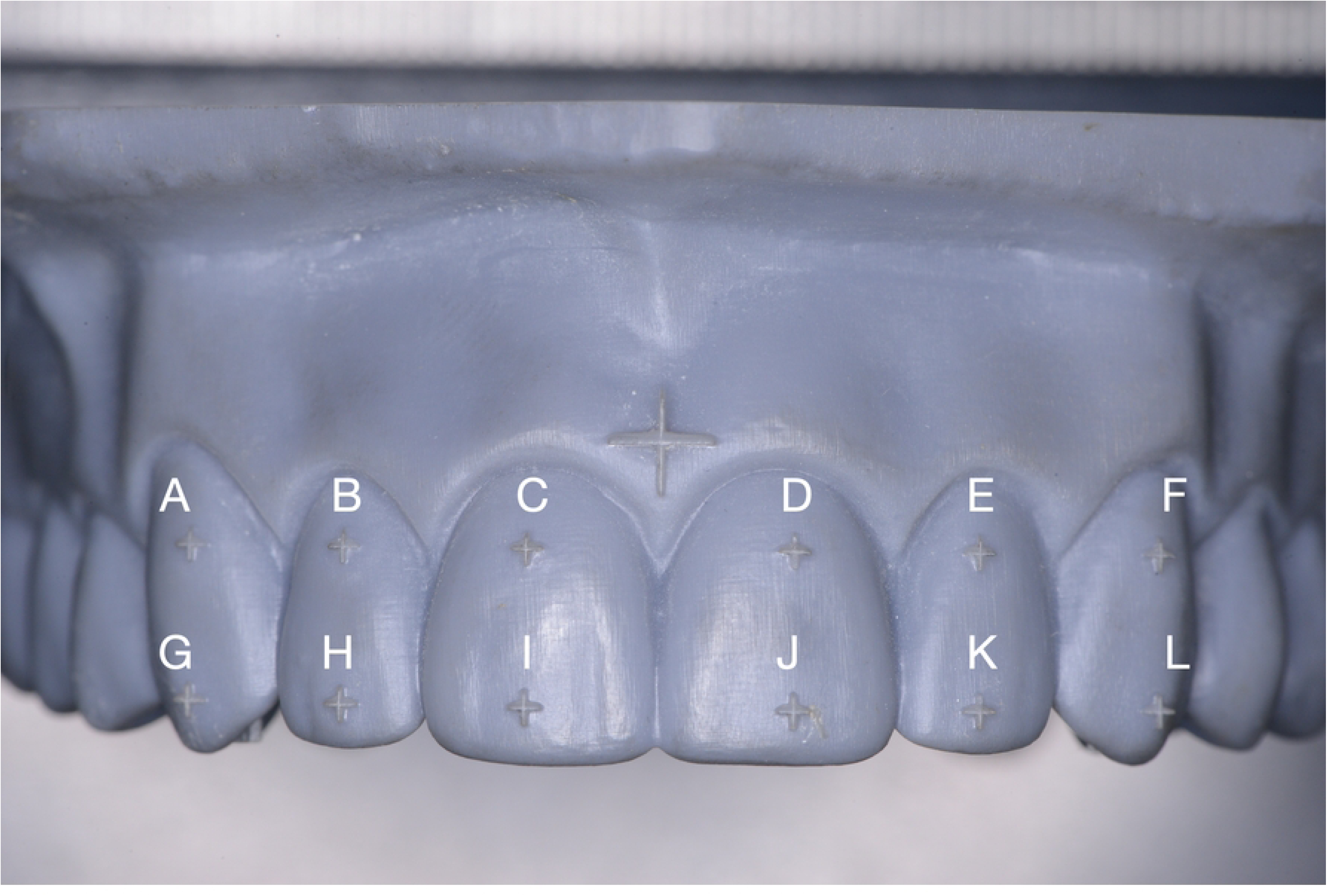
Model in frontal view with marked (A-L).

**Fig 9.**
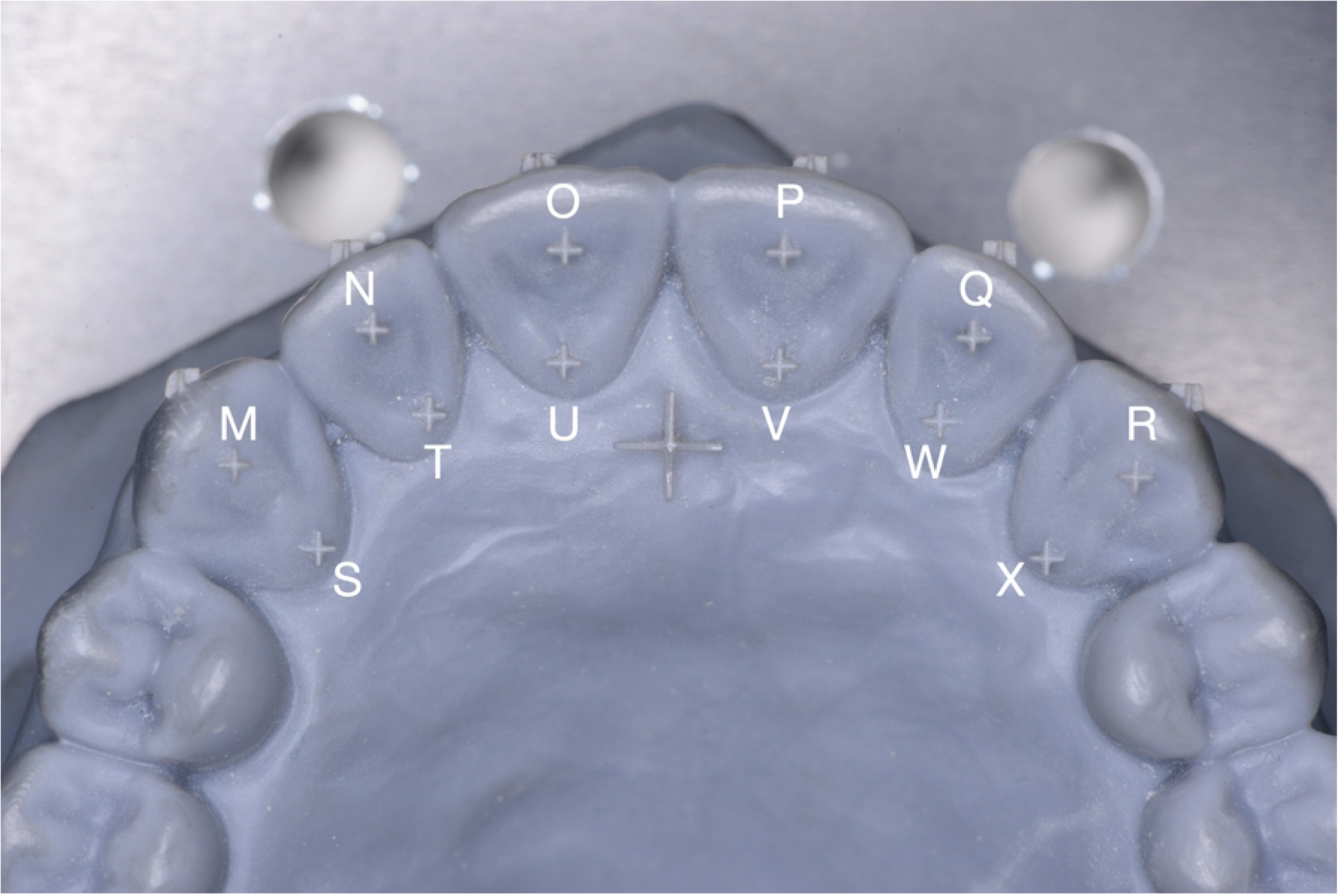
Model in occlusal view with marked (M-X).

### Model stand and bubble level setting

A custom model stand (A Best Engineering®, Thailand) was designed and fabricated from black aluminum alloy to securely hold the dental model. The stand allows the model to be fixed and precisely adjusted into both frontal and occlusal orientations at a 90-degree angle using a screw-lock mechanism. A linear ruler scale, marked in millimeters, was incorporated above the model position, with markings along both the horizontal and sagittal planes, to facilitate accurate distance calibration and secure positioning.

Adjustable components were integrated into the central portion of the stand, allowing fine-tuned movement in both the sagittal and horizontal planes. A vertical black aluminum pillar was constructed to support a smartphone holder, which was mounted onto a smartphone stabilizer (Rockbros®, China). This stabilizing system enabled controlled positioning of the smartphone relative to the model in multiple directions—sagittal, frontal, horizontal, and vertical—simultaneously. The integrated measurement scale further assisted in maintaining consistent positioning, as illustrated in Fig 10.

**Fig 10.**
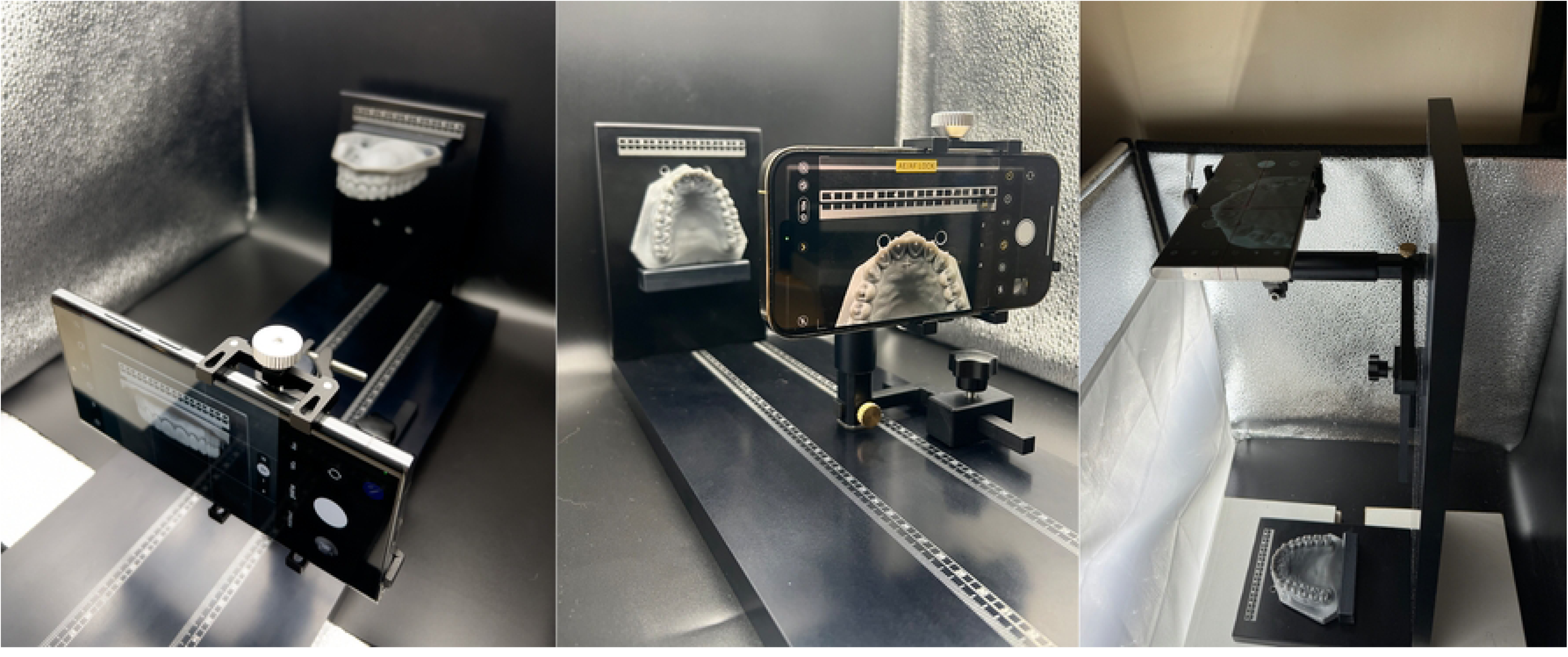
The model stand and smartphone stabilizer positioned inside the standardized light box.

Because the camera angle influences image distortion, all devices were positioned vertically above the model to maintain identical vertical and horizontal alignment. The 5 × 5 mm “+” marker positioned at the midline between the upper right and upper left central incisors served as the model’s central reference point for all photographs in both frontal and occlusal view. This marker was used to calibrate pixel dimensions and measure distances across images captured with different devices. To ensure consistency and minimize distortion, the central “+” marker was aligned precisely with the center of the photographic frame in all images.

For the control group, a DSLR camera equipped with a 105 mm f/2.8 macro lens and a ring flash was mounted on a vertical camera stand to ensure stable and consistent positioning. A bubble level was placed atop the camera body to verify vertical orientation, and the intersecting grid lines on the LCD screen were aligned precisely with the model’s central “+” marker in Fig 11.

**Fig 11.**
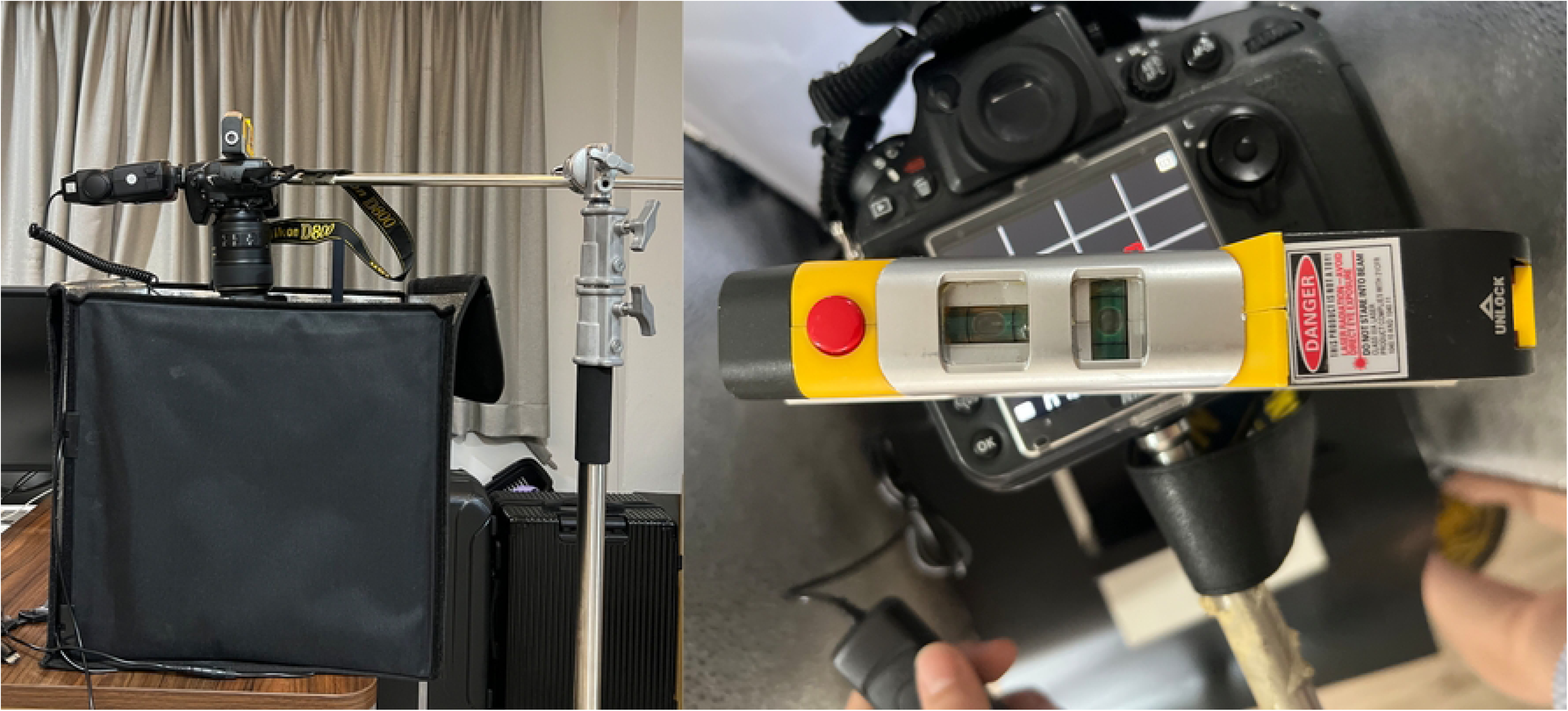
DSLR camera setup with bubble level alignment.

For the iPhone 15 Pro Max, the bubble level was displayed as a central “+” symbol on the camera interface. Alignment was achieved by superimposing this on-screen “+” with the 5 × 5 mm reference marker above the midline of the central incisors. In contrast, the Samsung Galaxy S23 Ultra displayed a circular bubble level indicator. To standardize alignment with the iPhone, two red threads were arranged to form a crosshair over the circular bubble level, enabling accurate superimposition with the model’s central reference point in Fig 12.

**Fig 12.**
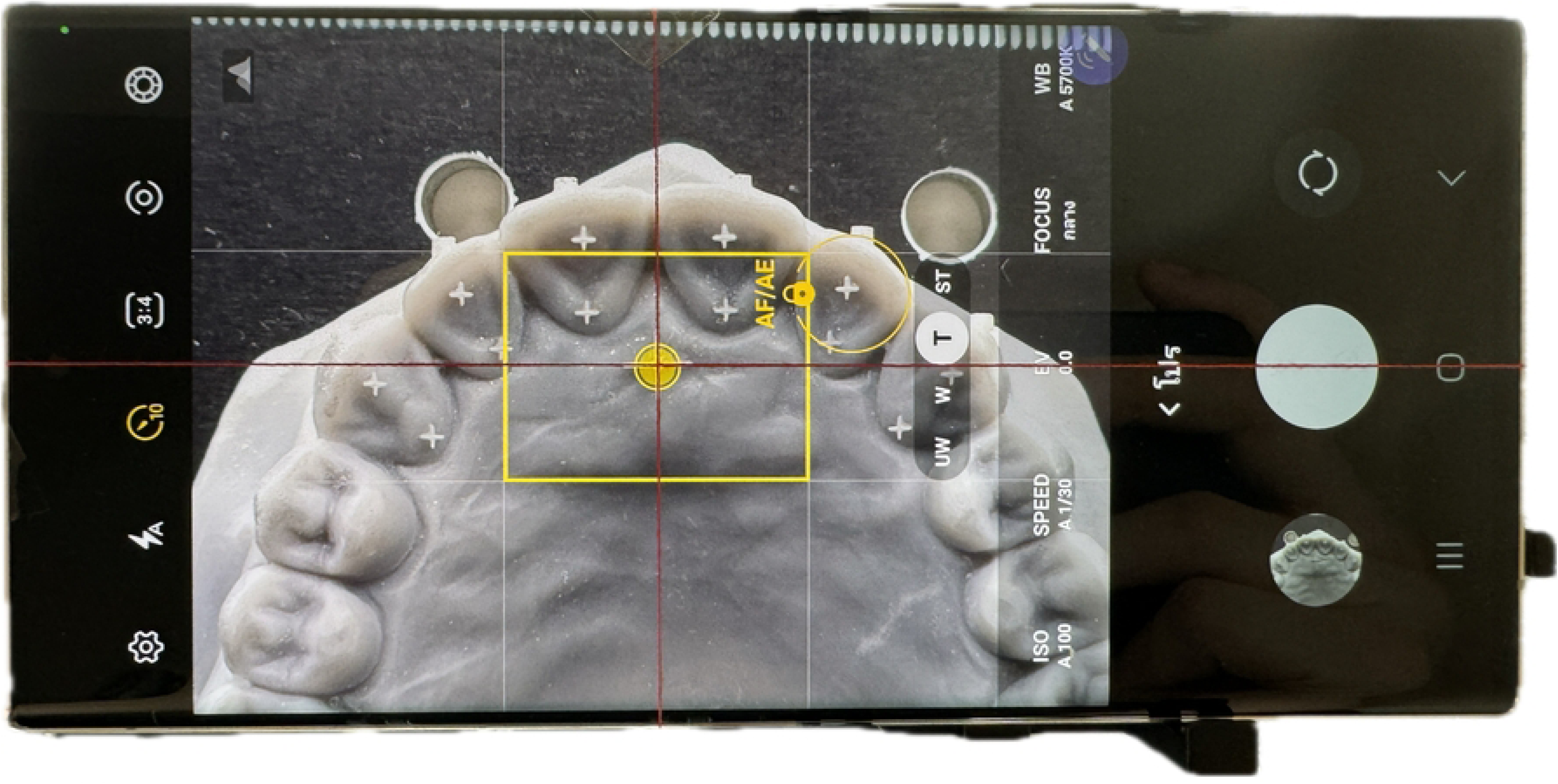
Bubble level alignment configuration for the Samsung Galaxy S23 Ultra.

### Environmental Conditions

All photographs were captured in a closed room under controlled lighting conditions using a unidirectional LED light source housed within a standardized light box (Puluz®, China). For the DSLR camera, the high-aperture setting (F32) produced underexposed images; therefore, a ring flash (Godox ML-150, Godox®, China) was employed as the primary light source. However, the use of a ring flash resulted in minor variations in color tone when compared with the smartphone images captured solely under LED illumination. All smartphone devices were operated in automatic mode to maintain consistent exposure and focus. The specifications of cameras, lenses, and lighting devices are summarized in Table 2.

### Examiner and camera standardization

All images were captured by a single operator to ensure procedural consistency. Linear measurements of tooth dimensions were also performed by the same examiner to minimize inter-examiner variability.

Reproducibility of photographic acquisition was ensured by strictly standardizing all imaging parameters, as summarized in Table 3. Each device captured images in a 4:3 aspect ratio. Both smartphones were mounted on the stabilizer system attached to the model stand, while the DSLR camera was fixed on a vertical stand.

**Table 3.**
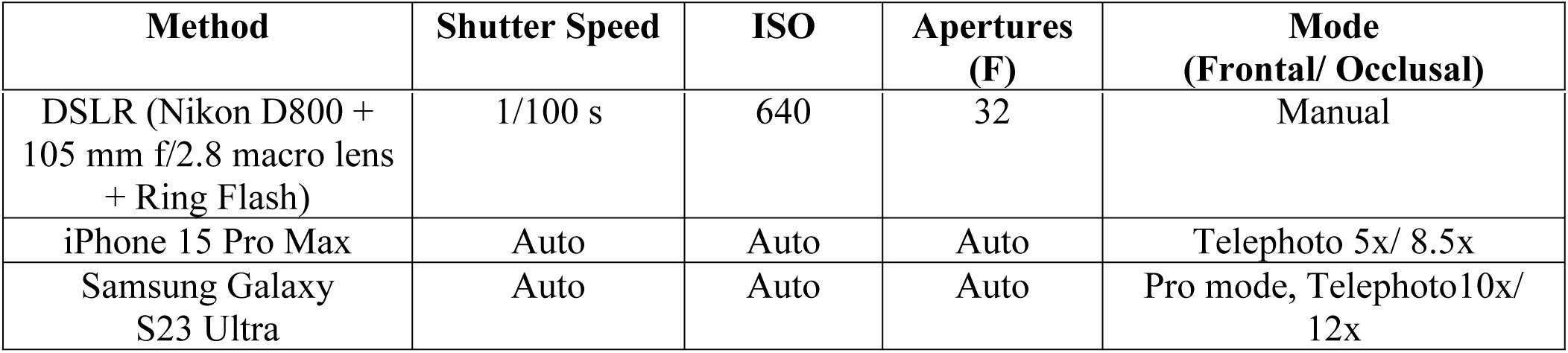
Demonstrates the standard cameras setting parameters.

The distance between the camera lens and the dental model was standardized at 35 cm for both frontal and occlusal views to ensure reproducibility across all image sets. The focus point was consistently positioned at the center of tooth 12. For smartphone cameras, the auto exposure/autofocus (AE/AF) lock was engaged to prevent focus drift, while the DSLR camera employed manual focus to maintain consistent focal control on the same reference area. The area of interest was restricted to the anterior segment (canine to canine), corresponding to the typical esthetic zone routinely emphasized in clinical dental photography.

### DSLR photography

The DSLR camera was equipped with a 105 mm f/2.8 macro lens and a ring flash, served as the control device. All exposure parameters followed Table 3. Ten frontal-view and ten occlusal-view images were acquired, totaling twenty DSLR photographs for analysis.

A 10-second timer and wired remote shutter minimized vibration during exposure. Any blurred or misfocused images were immediately retaken under identical conditions to maintain consistency.

### Smartphone photography

For the iPhone 15 Pro Max, the camera-to-model distance remained fixed. The device operated in Telephoto mode, with AE/AF locked on tooth 12. A 5× zoom was used for ten frontal-view images, and an 8.5× zoom for ten occlusal-view images as Fig 13. Although the iPhone’s telephoto lens provides up to 5× optical zoom, further magnification employed digital zoom, which may reduce resolution and sharpness. Due to a system limitation, the iPhone 15 Pro Max automatically activates burst mode when the 10-second timer is used, leading to reduced image resolution; therefore, photographs with this device were captured manually. A 10-second interval was observed between each capture, and the smartphone remained fixed on the stabilizer.

**Fig 13.**
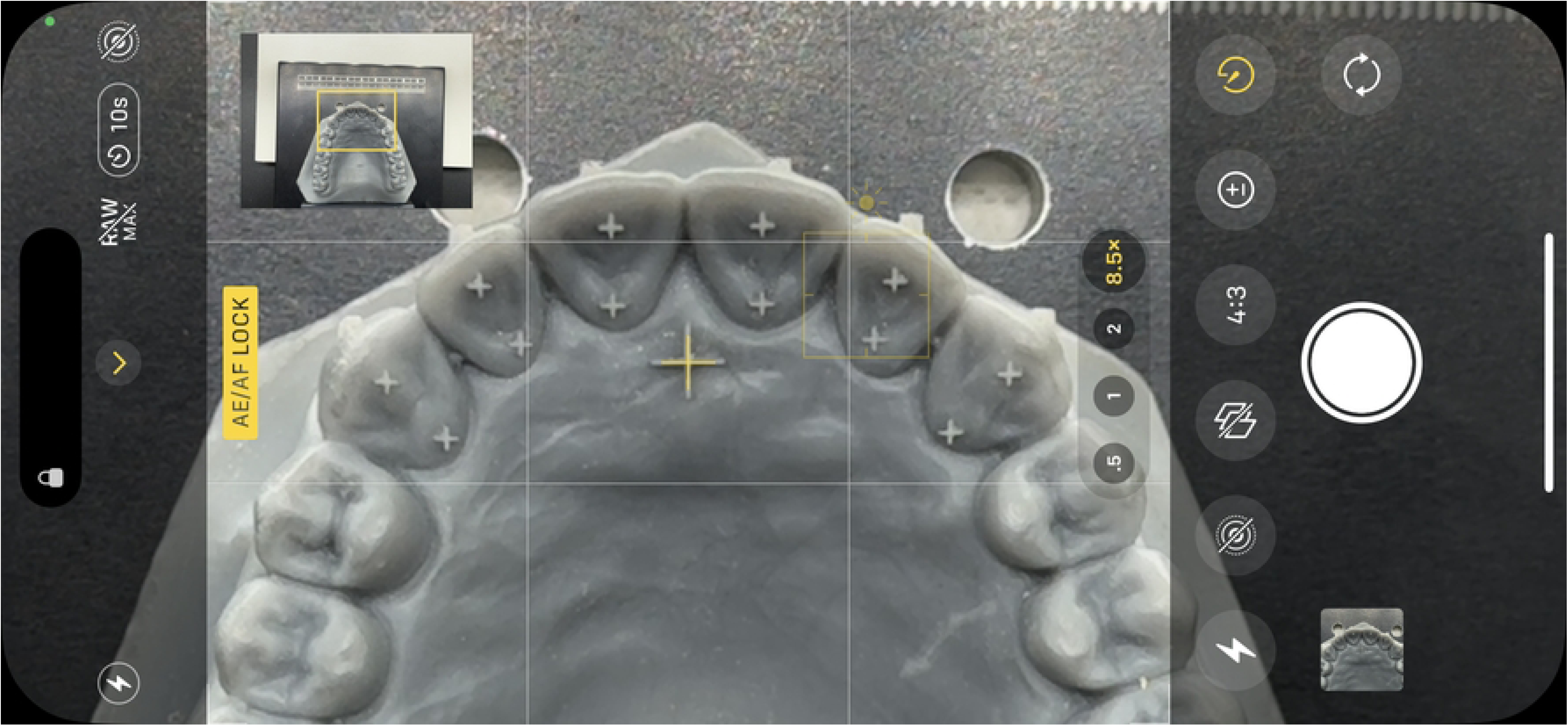
Camera interface of the Apple iPhone 15 Pro Max in Telephoto mode.

The Samsung Galaxy S23 Ultra was operated in Pro camera mode with the Telephoto lens and AE/AF lock enabled. A 10× zoom was used for frontal images and 12× zoom for occlusal images, yielding twenty photographs in total. The S Pen stylus served as a wireless remote shutter to prevent motion-induced blur. Although the Samsung Galaxy S23 Ultra telephoto lens provides up to 10× optical zoom, any magnification beyond this level relies on digital zoom, which may consequently reduce image resolution and sharpness.

Upon completion, all 60 image files were organized into three device-specific folders. No post-processing, cropping, or image enhancement was performed prior to measurement as illustrated in Figs 14 and 15.

**Fig 14.**
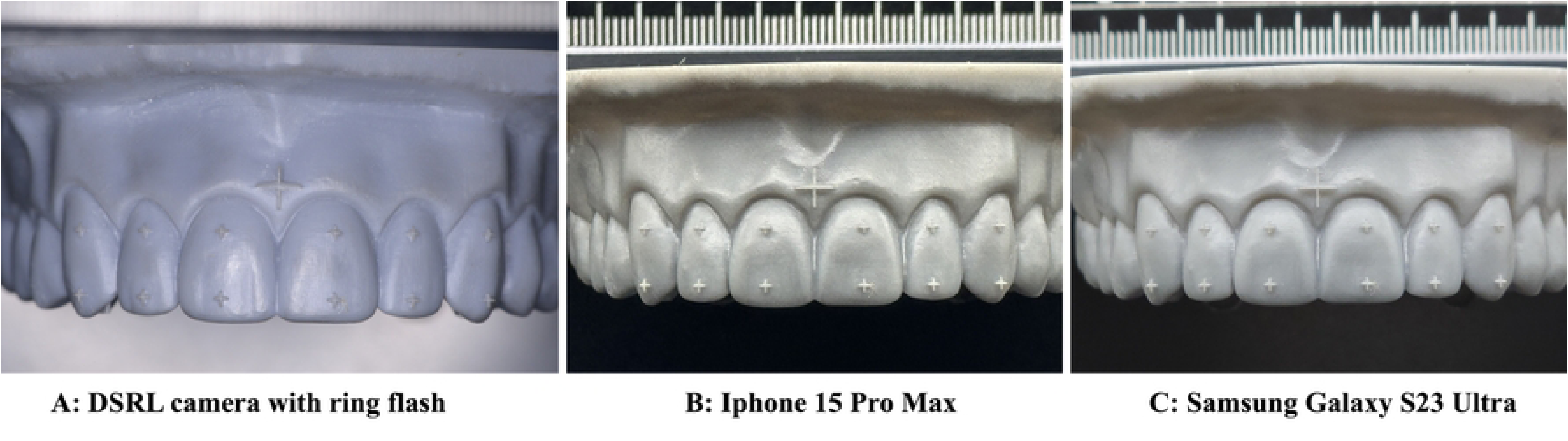
(A,B,C) Frontal view photographs of the dental model.

**Fig 15.**
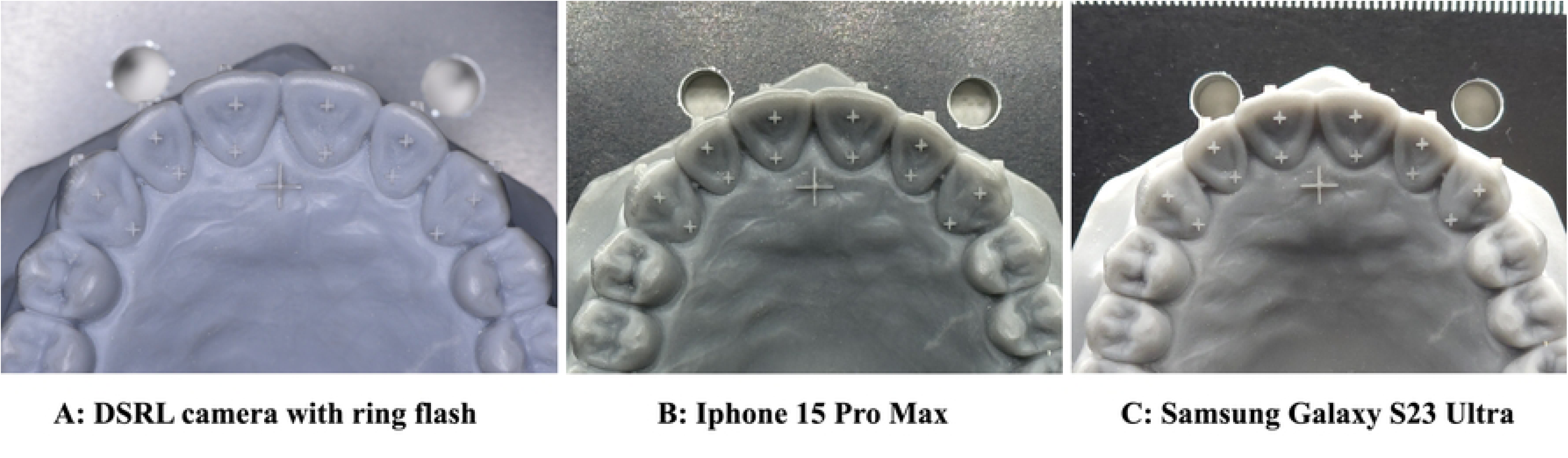
(A,B,C) Occlusal view photographs of the dental model.

### Linear dimensional measurements

Image analysis was performed using ImageJ software (as shown in Fig 16) to measure the linear dimensions of the teeth in MacBook version. The central reference “+” marker, measuring 5 × 5 mm and positioned between upper right central incisor and upper left central incisor, was used to calibrate the scale in millimeters for each photograph. This calibration step was conducted separately for images from each device prior to performing any additional linear measurements.

**Fig 16.**
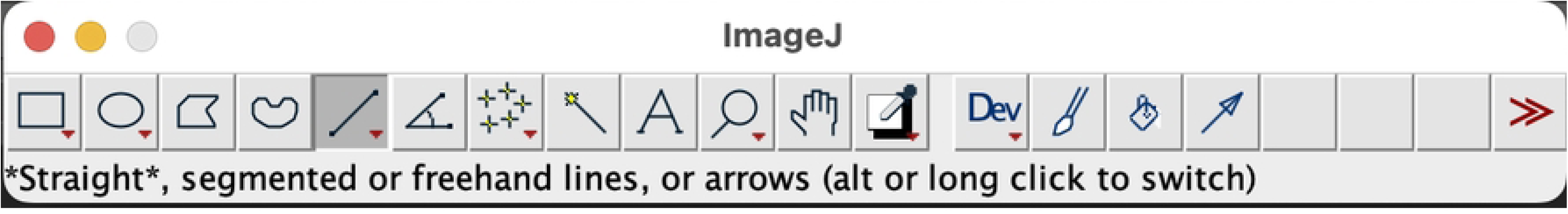
ImageJ software.

To enhance measurement accuracy, each linear dimension was measured three times, and the average value was used for analysis. For the central reference “+” marker known as “Control 1” in the frontal view image and “Control 2” in the occlusal view image, calibration was conducted in three directions—left to right, right to left, and top to bottom—to verify the consistency of examiner measurements. ImageJ software was utilized for image analysis, with yellow point-to-point lines applied to set the scale for calibration and perform linear measurements as illustrated in Fig 17.

**Fig 17.**
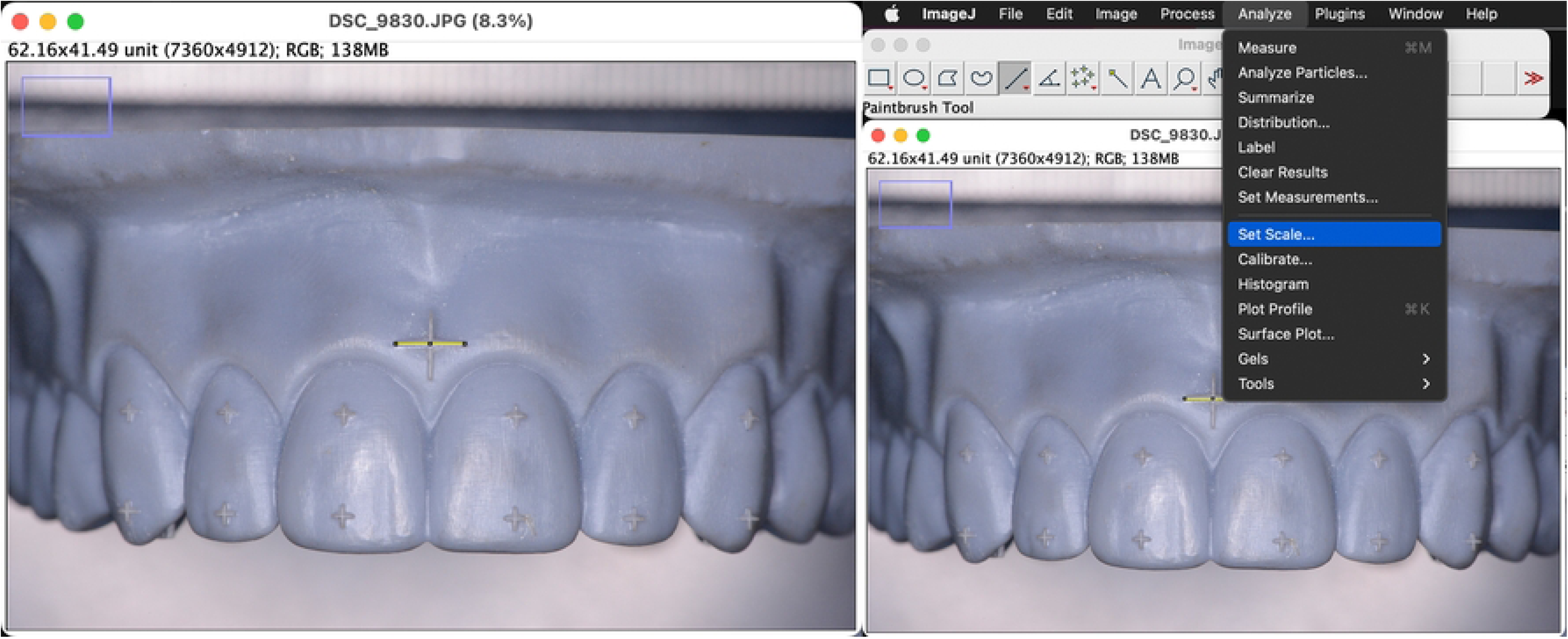
ImageJ software with setting scale for calibration.

The “Control 1” and “Control 2” markers, used as a reference, were scaled to calibrate pixel-to-millimeter conversion for each image across different devices, allowing for cross-device comparison of results (Fig 18). Linear distances between defined points were measured using a yellow reference line.

**Fig 18.**
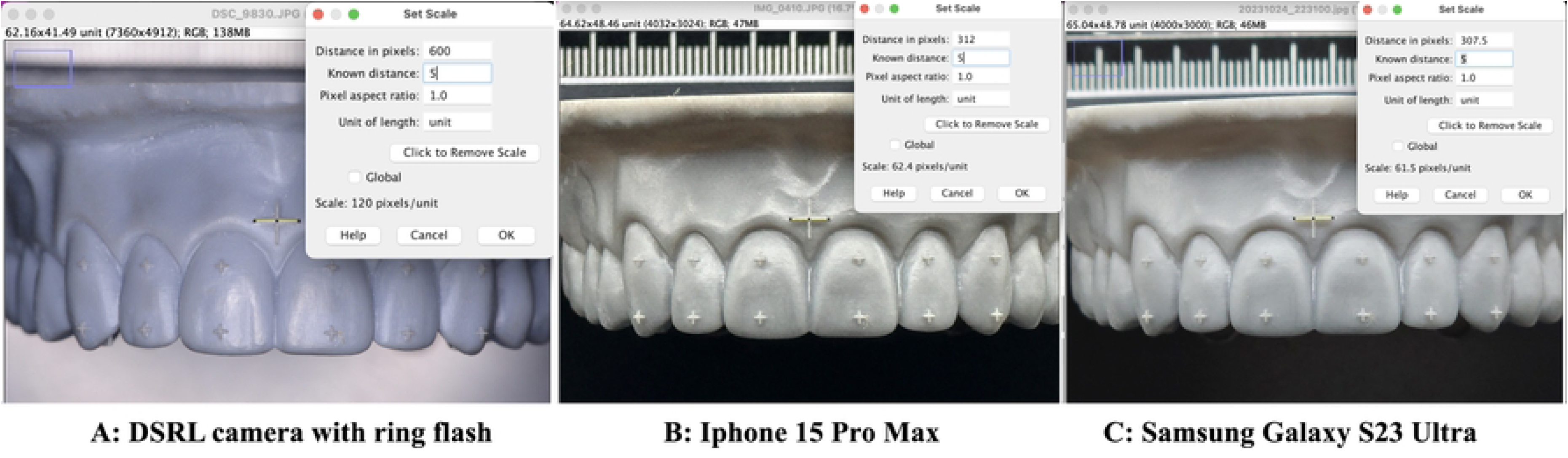
(A,B,C) Calibrate the pixel of photograph in ImageJ software.

The measurement values were recorded and presented in terms of “length” in the results table. Each measurement was repeated three times in the specified order, and the resulting data were compiled and presented, as shown in Figs 18 and 19.

**Fig 19.**
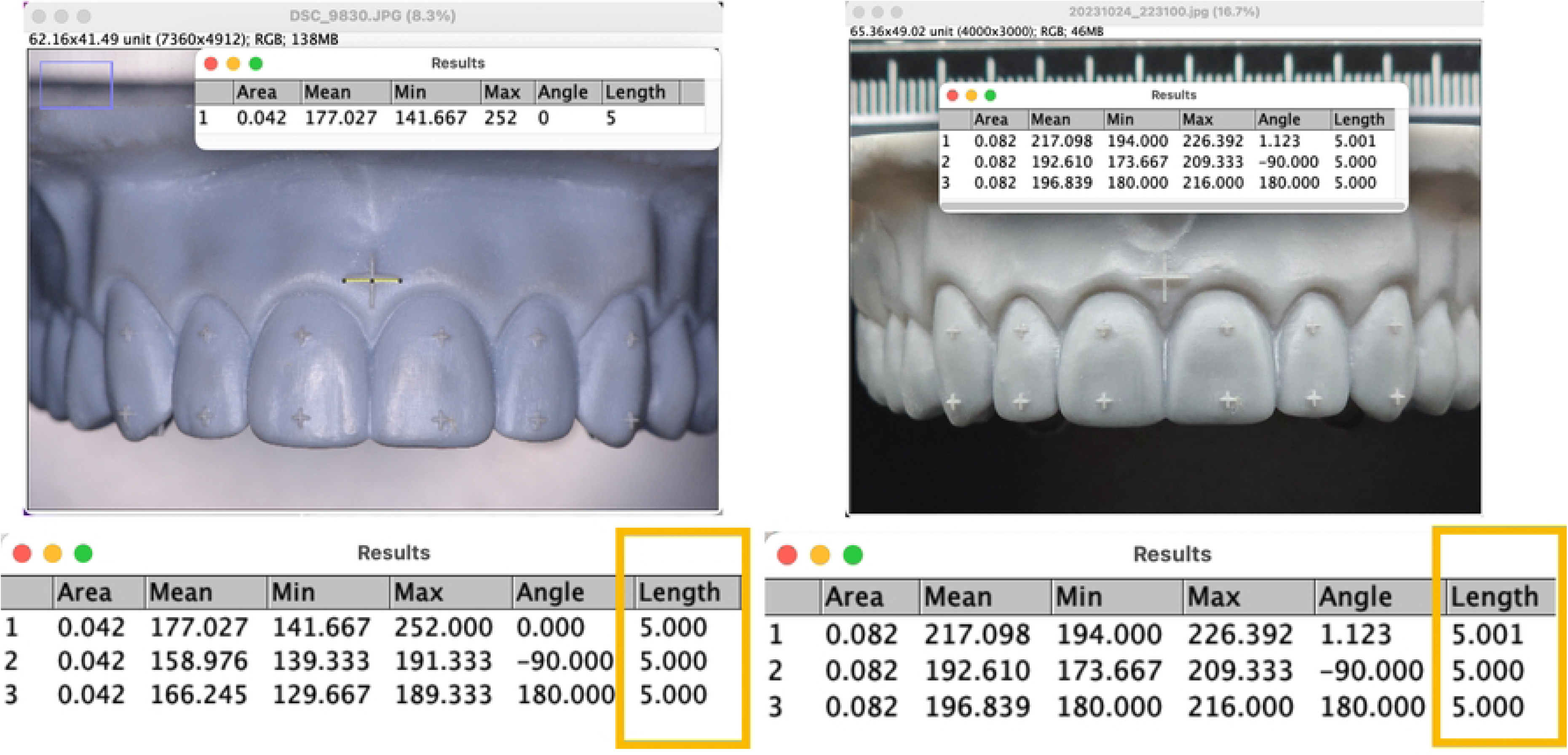
Example of result table of calibration in DSLR and smartphone.

Following calibration of the photographs from each device using the reference markers “Control 1” for frontal view images and “Control 2” for occlusal view images, linear dental measurements were conducted between the cross-sections of the “+” markers positioned at the incisal third and cervical third on the labial surfaces of the upper right and left canines, lateral incisors, and central incisors.

Each measurement was performed three times between corresponding points, and the average value was used for analysis. Measurements were obtained from both frontal and occlusal perspectives to ensure a comprehensive dimensional analysis.

Linear measurements were obtained from the “+” markers positioned at the incisal third and cervical third of the upper anterior teeth. In the frontal view, the distances between the upper right and left central incisors (teeth 11–21) were measured along the lines from point C to D and I to J. Similarly, measurements for the upper lateral incisors (teeth 12–22) were recorded along lines from point B to E and H to K, and for the upper canines (teeth 13–23), along lines from point A to F and G to L. All linear measurements were recorded in millimeters.

In the occlusal view, following the calibration with “Control 2”, corresponding linear measurements were taken between the “+” markers positioned at the incisal and cervical thirds of the respective teeth. Specifically, measurements for teeth 11–21 were recorded along lines O to P and U to V; for teeth 12–22, along lines N to Q and T to W; and for teeth 13–23, along lines M to R and S to X. Each measurement was performed three times, and the average value was calculated to ensure accuracy for subsequent analysis, as illustrated in Fig 20.

**Fig 20.**
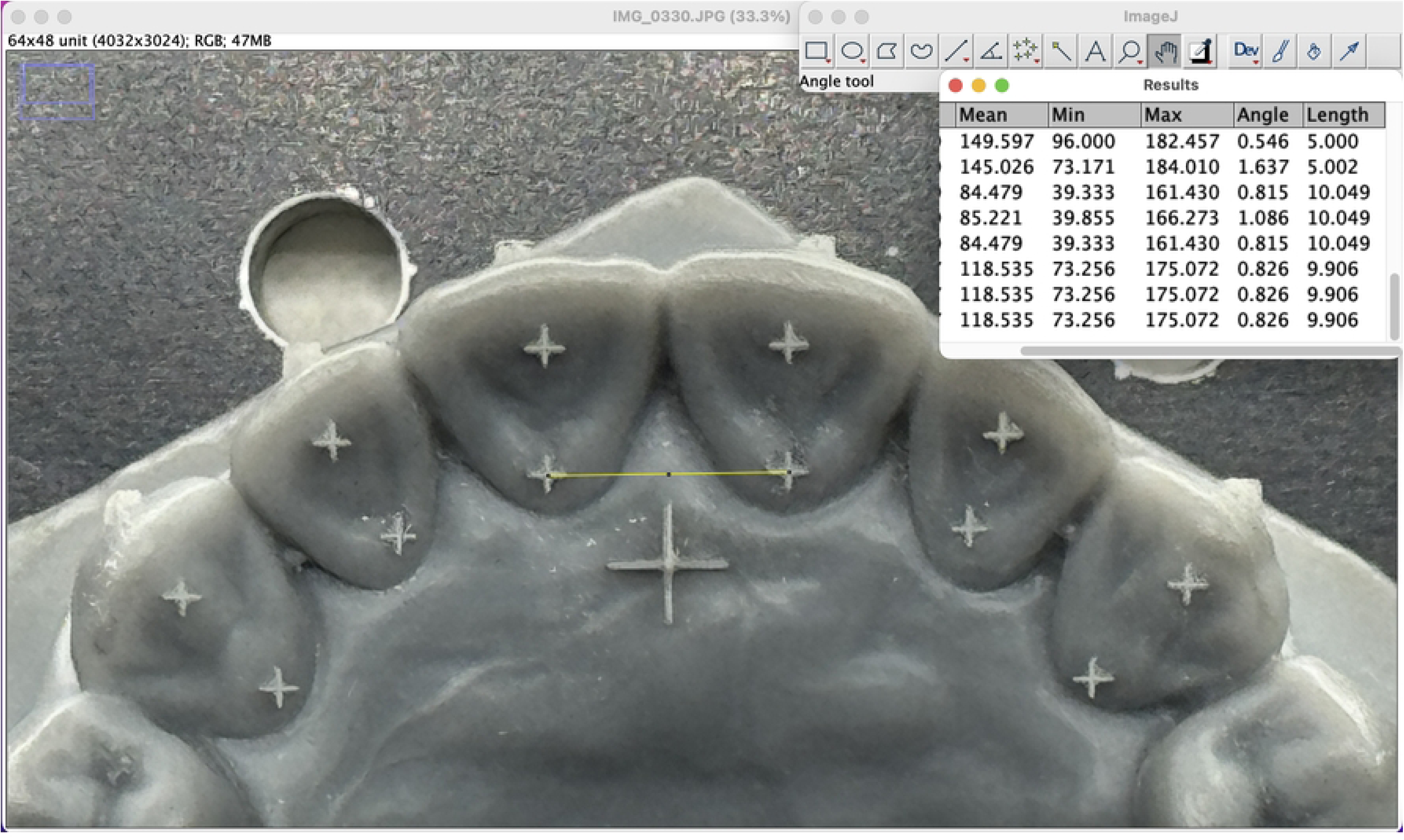
Example of the result in occlusal view image.

The ImageJ software generated linear dimensional measurements for teeth 11–21, 12–22, and 13–23 in both frontal and occlusal views. For each device, 10 frontal view images (30 images in total) and 10 occlusal view images (30 images in total), 60 images were analyzed. The linear distances, measured in millimeters from defined point to point, were recorded in the results table. Each measurement was repeated three times, and the average value was calculated using Microsoft Excel to ensure accuracy before being used for numerical comparison across methods.

## 5. Result

Descriptive statistics was calculated using SPSS (Statistical Package for the Social Sciences) version 30 (IBM, Chicago, IL, USA). The numerical data representing linear tooth dimensions obtained from the three different imaging methods were recorded and analyzed. The measurements, expressed as “length,” were taken from the calibrated results tables, which are presented as mean and standard deviation (SD) in Tables 4 and 5. Given the sample size of 10 per group (n=10), the accuracy of each method was compared to the control group (DSLR camera) using Friedman’s test with Bonferroni correction for multiple comparisons. The results were considered statistically significant at the 0.05 level (p < 0.05).

**Table 4.** Result of measurement in frontal view.

| Frontal view | Nikon D800<br>*Mean (SD)* | iPhone 15 Pro Max<br>*Mean (SD)* | Samsung Galaxy S23 Ultra<br>*Mean (SD)* |
| --- | --- | --- | --- |
| Control 1 | 5 (0.000) | 5 (0.000) | 5 (0.000) |
| C_D | 12.70 (0.007) | 13.137 (0.033) | 12.821 (0.073) |
| I_J | 12.700 (0.007) | 13.138 (0.003) | 12.84 (0.007) |
| B_E | 29.890 (0.020) | 30.982 (0.000) | 30.293 (0.013) |
| H_K | 30.069 (0.002) | 31.102 (0.031) | 30.396 (0.003) |
| A_F | 45.602 (0.011) | 47.326 (0.029) | 45.972 (0.017) |
| G_L | 45.611 (0.000) | 47.254 (0.000) | 45.984 (0.000) |

**Table 5.** Result of measurement in occlusal view.

| Occlusal view | Nikon D800<br>*Mean (SD)* | iPhone 15 Pro Max<br>*Mean (SD)* | Samsung Galaxy S23 Ultra<br>*Mean (SD)* |
| --- | --- | --- | --- |
| Control 2 | 5 (0.000) | 5 (0.000) | 5 (0.000) |
| O_P | 10.065 (0.007) | 10.282 (0.031) | 10.149 (0.027) |
| U_V | 9.914 (0.219) | 10.197 (0.001) | 10.087 (0.032) |
| N_Q | 27.535 (0.001) | 28.270 (0.046) | 27.979 (0.042) |
| T_W | 23.468 (0.000) | 24.10 (0.041) | 23.836 (0.048) |
| M_R | 41.368 (0.015) | 42.454 (0.000) | 41.818 (0.042) |
| S_X | 33.47 (0.001) | 34.316 (0.000) | 33.898 (0.000) |

Friedman’s test revealed no statistically significant difference among the control measurements across all devices, confirming consistent baseline calibration. However, significant differences were detected in multiple inter-point measurements obtained using the iPhone 15 Pro Max when compared with the DSLR camera, indicating measurable linear dimensional shifts. In contrast, the Samsung Galaxy S23 Ultra demonstrated no statistically significant differences from the DSLR camera across most measured parameters, suggesting dimensional consistency.

Accordingly, the null hypothesis was retained for the Samsung Galaxy S23 Ultra, signifying no significant difference from the DSLR reference in image proportions. Conversely, the null hypothesis was rejected for the iPhone 15 Pro Max, indicating a significant difference in the linear dimensional proportions compared with the DSLR camera.

These findings suggest that while the iPhone 15 Pro Max tends to overestimate linear dimensions, the Samsung Galaxy S23 Ultra provides measurements closely aligned with the DSLR, showing greater dimensional accuracy.

The comparison of the three different devices was demonstrated in graphs, showing the linear measurements in the frontal view for each point-to-point distance between the “+” markers positioned at the incisal third and cervical third on the labial surfaces of the upper right and left canines, lateral incisors, and central incisors, as illustrated in Figs 21–27. Similarly, the results from the occlusal view images are presented in Figs 28–34.

**Fig 21.**
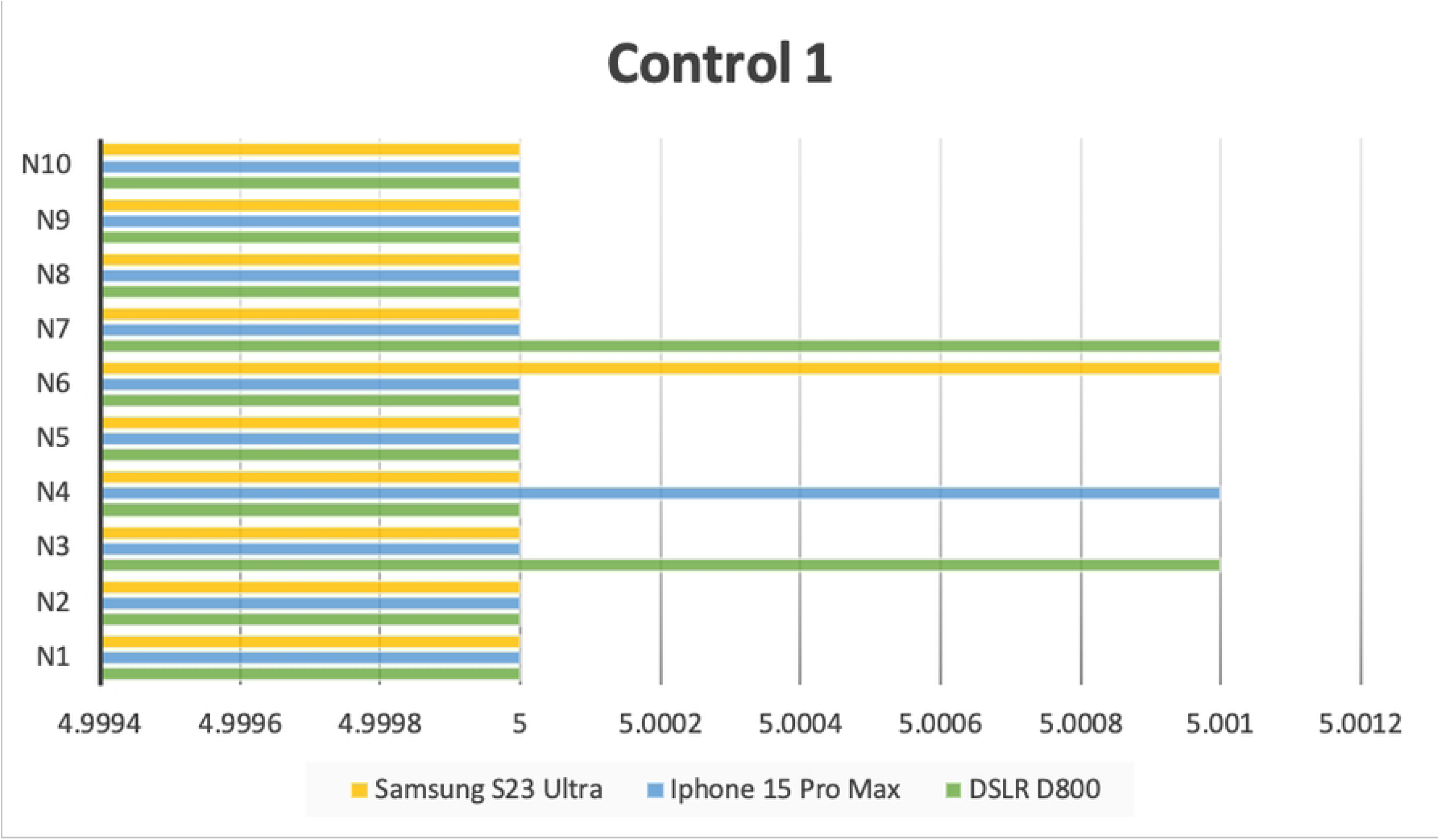
Result of Control1 in frontal view image.

**Fig 22.**
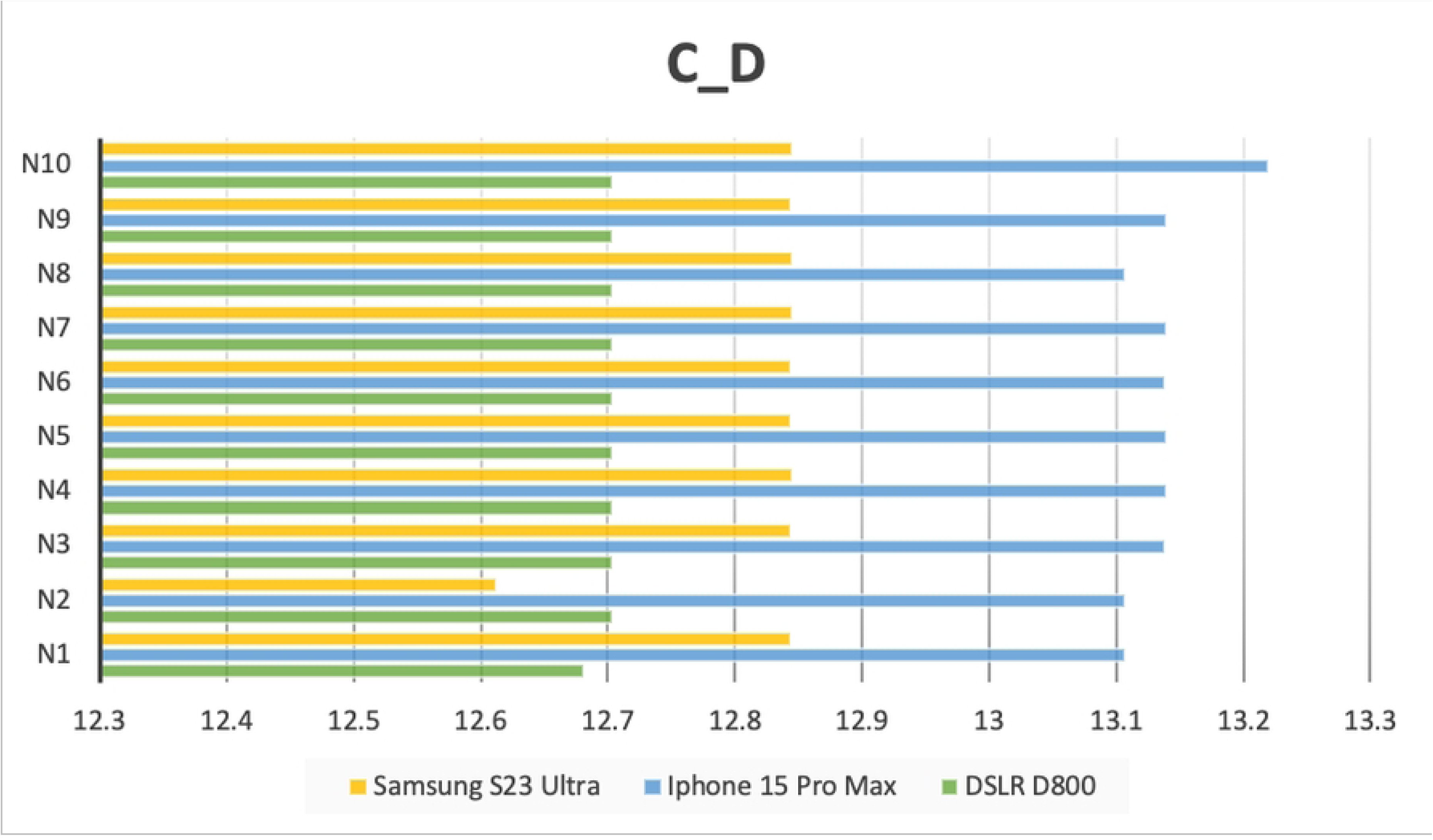
Result of C_D in frontal view image.

**Fig 23.**
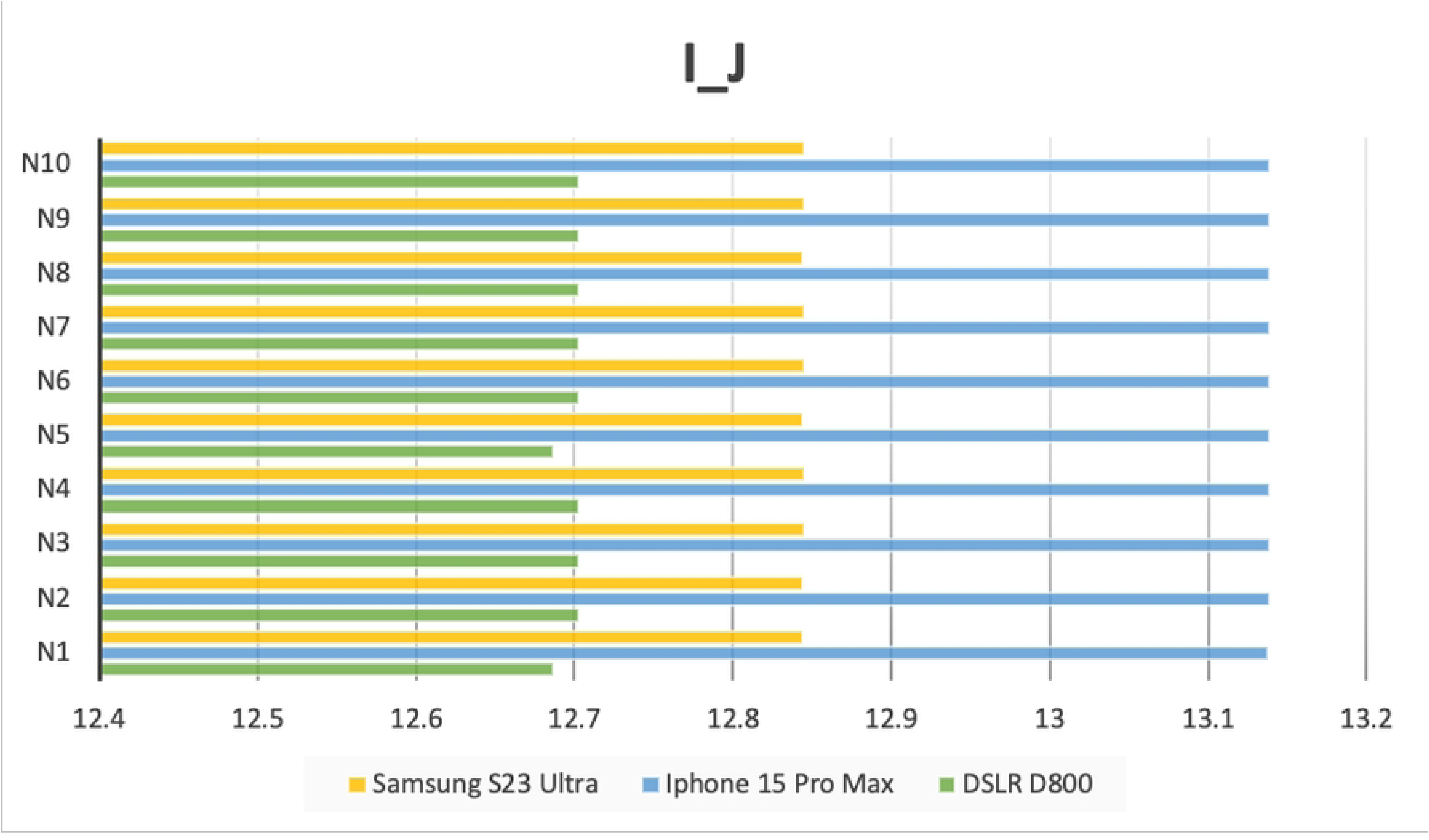
Result of I_J in frontal view image.

**Fig 24.**
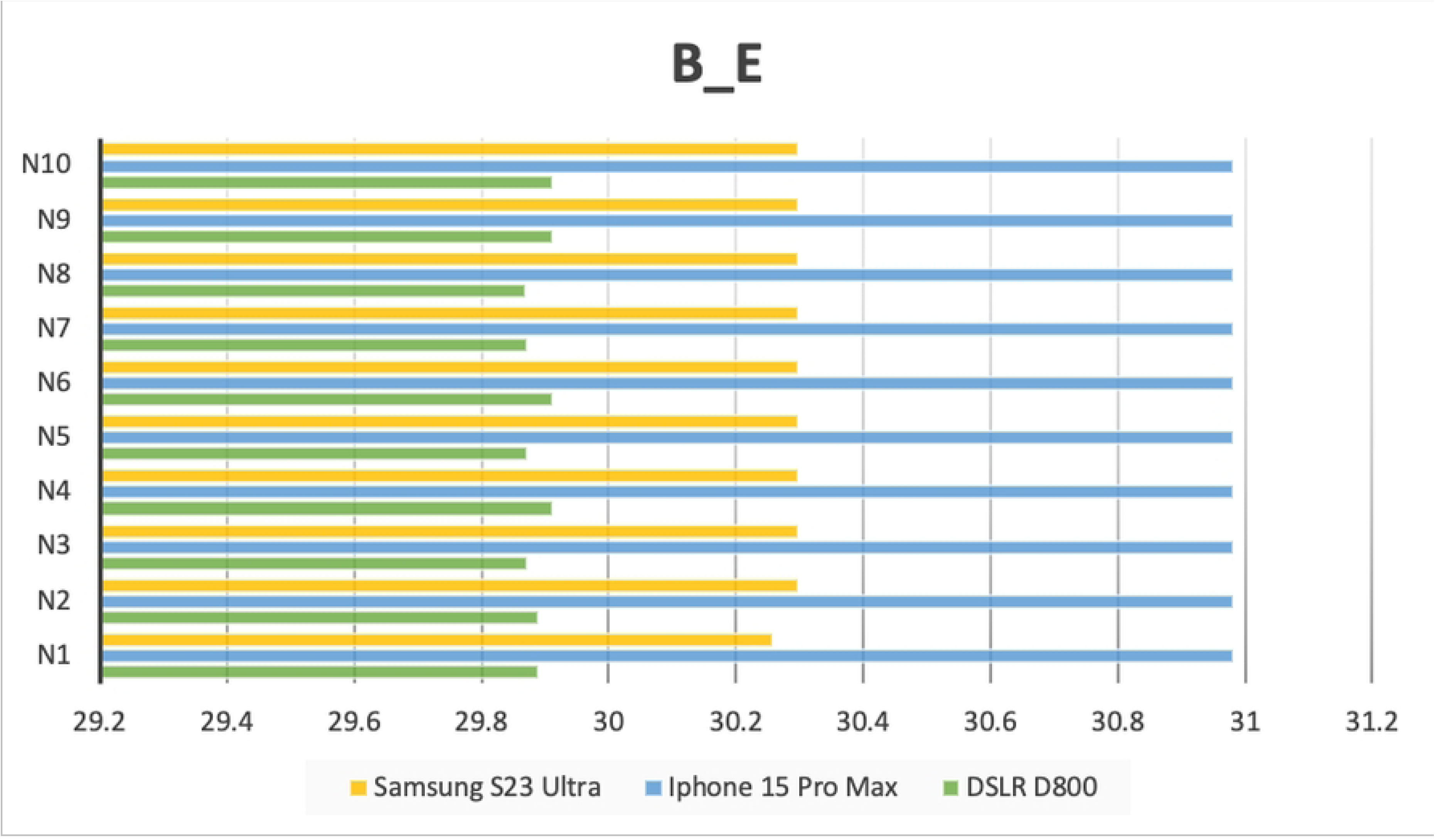
Result of B_E in frontal view image.

**Fig 25.**
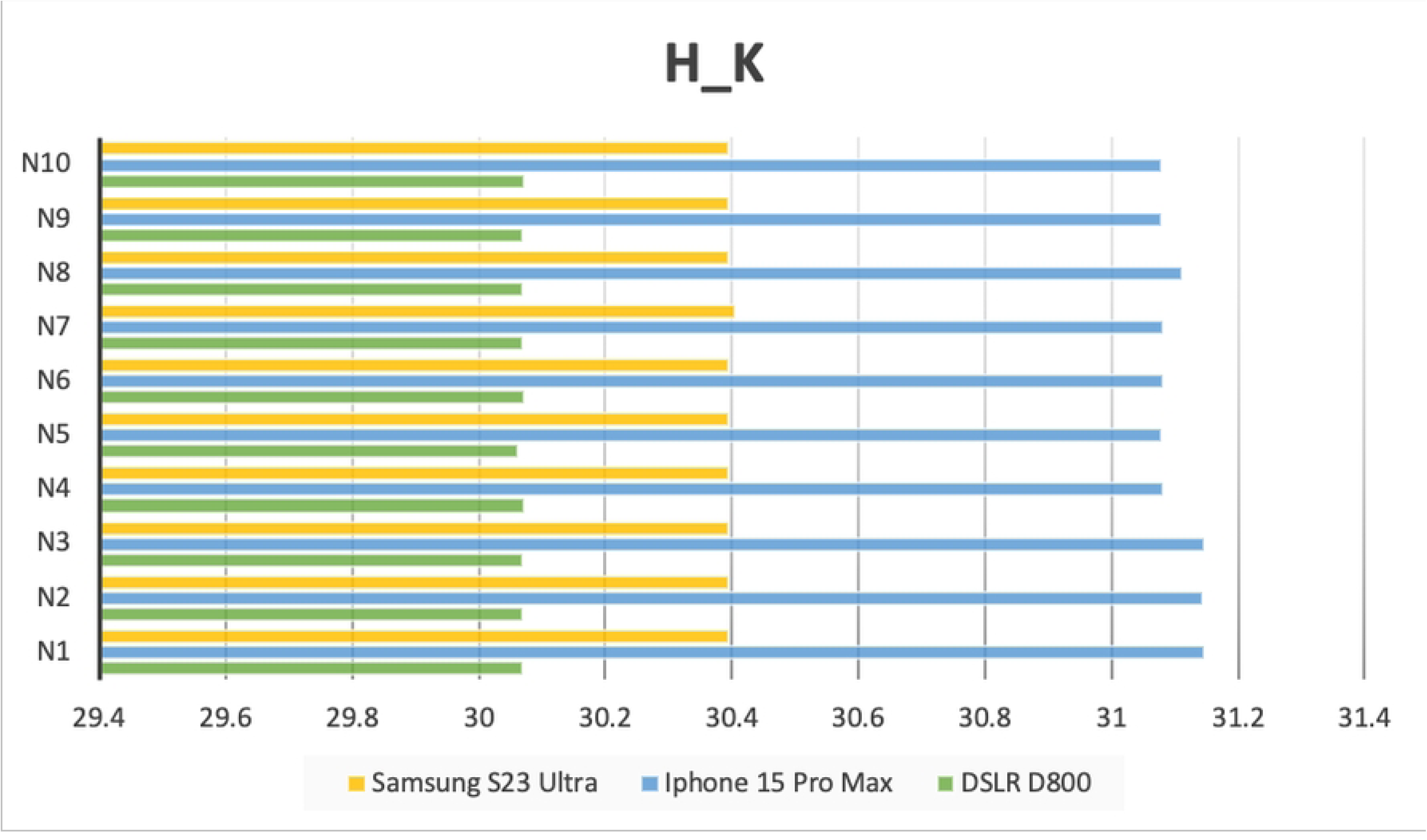
Result of H_K in frontal view image.

**Fig 26.**
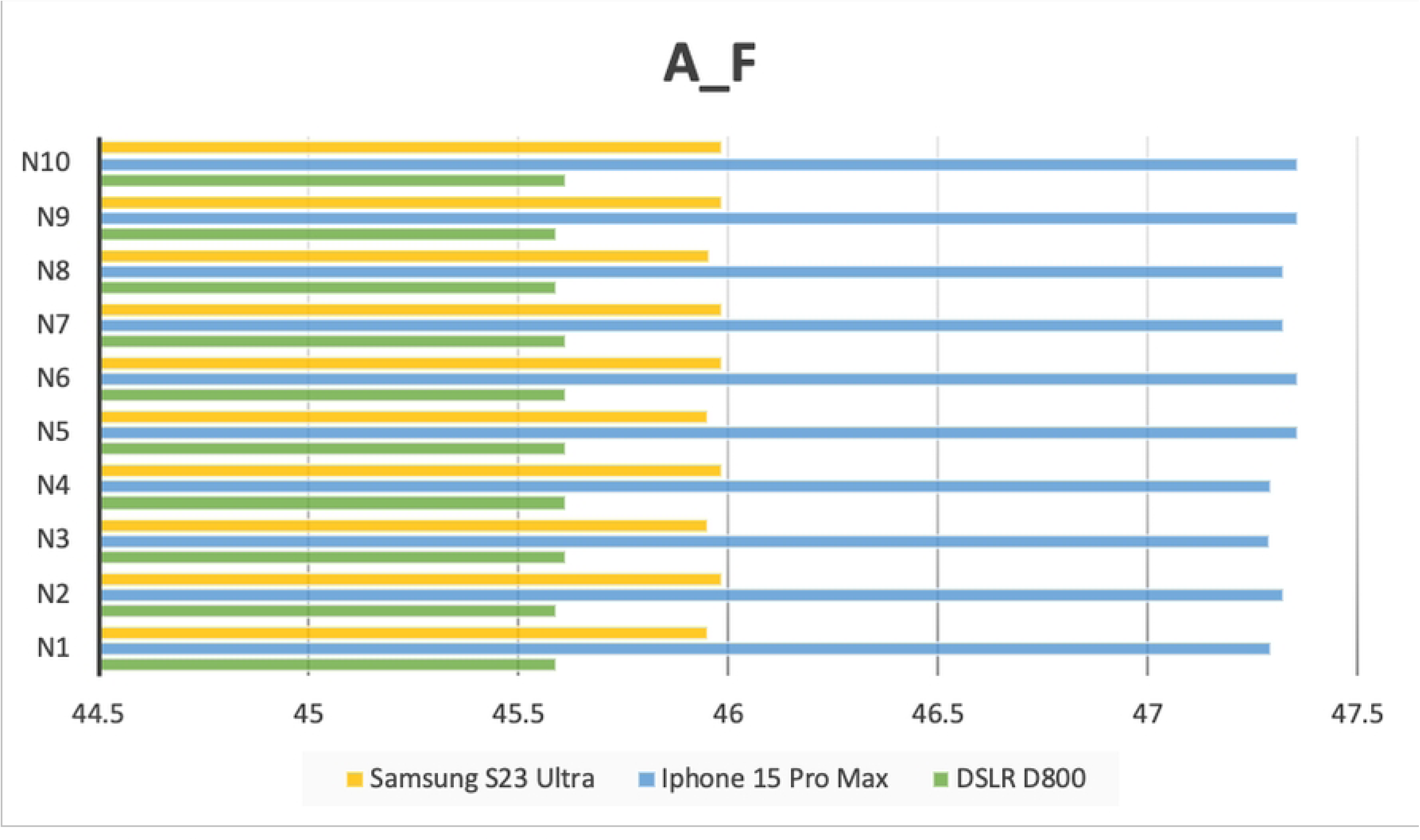
Result of A_F in frontal view image.

**Fig 27.**
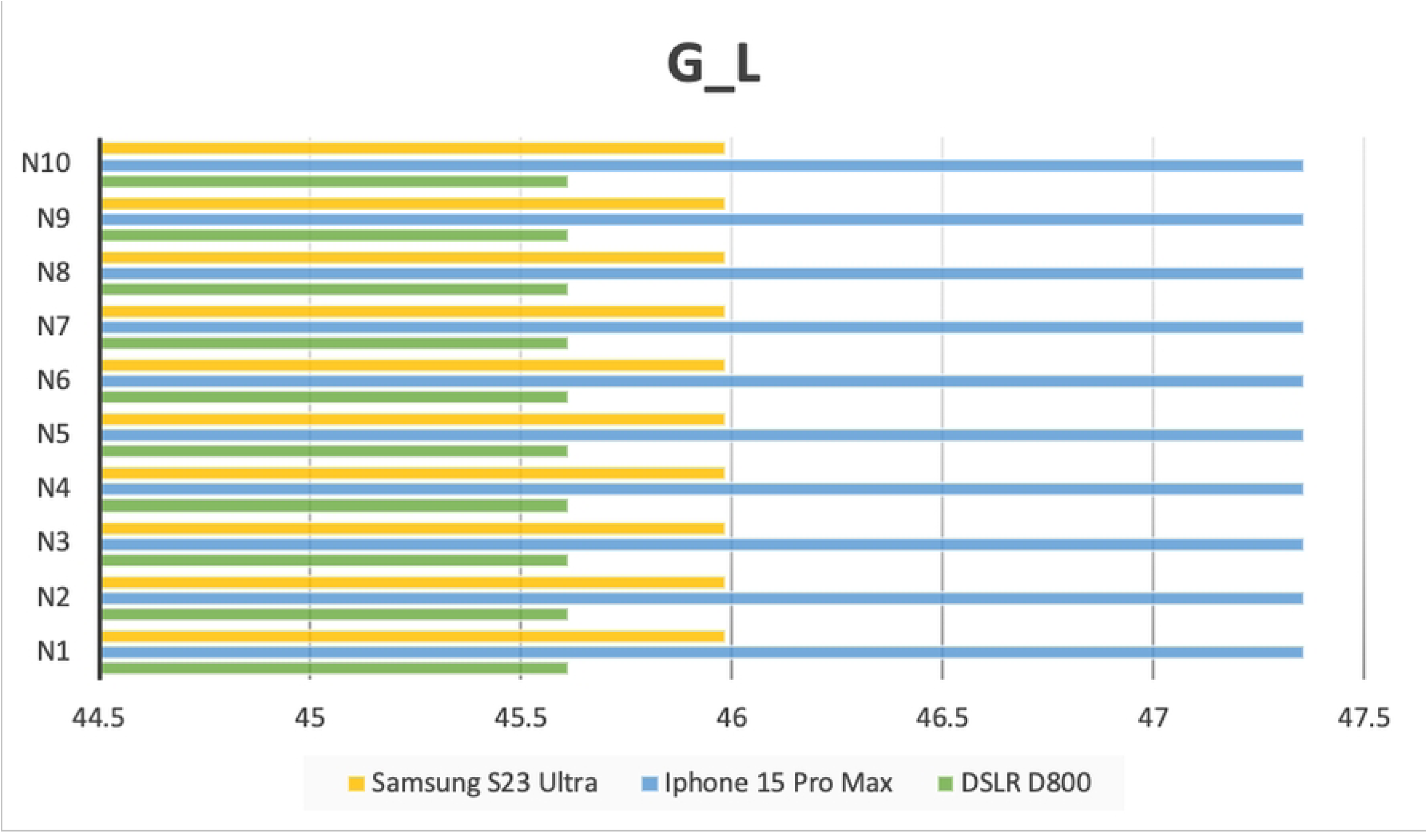
Result of G_L in frontal view image.

**Fig 28.**
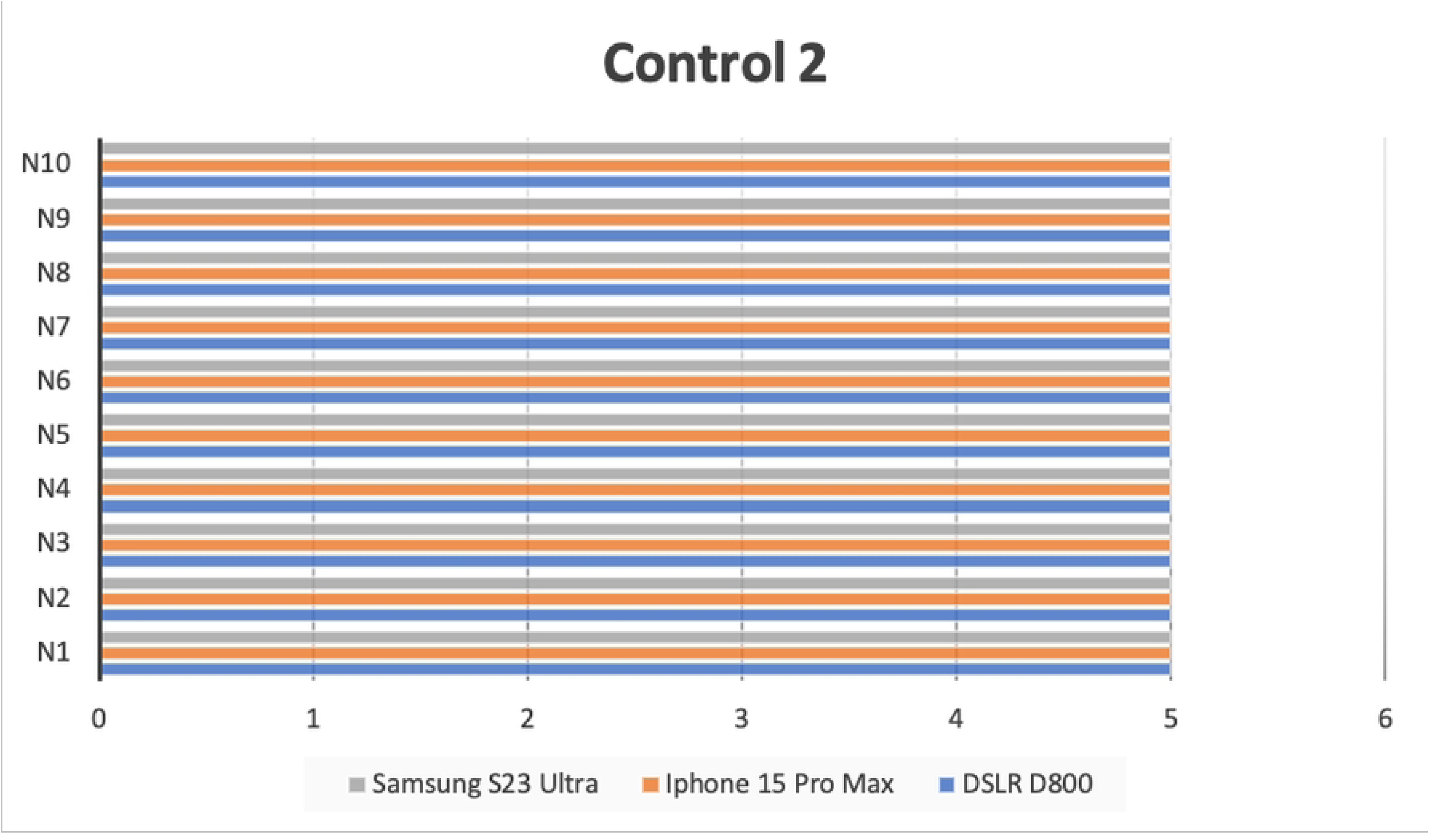
Result of Control2 in occlusal view image.

**Fig 29.**
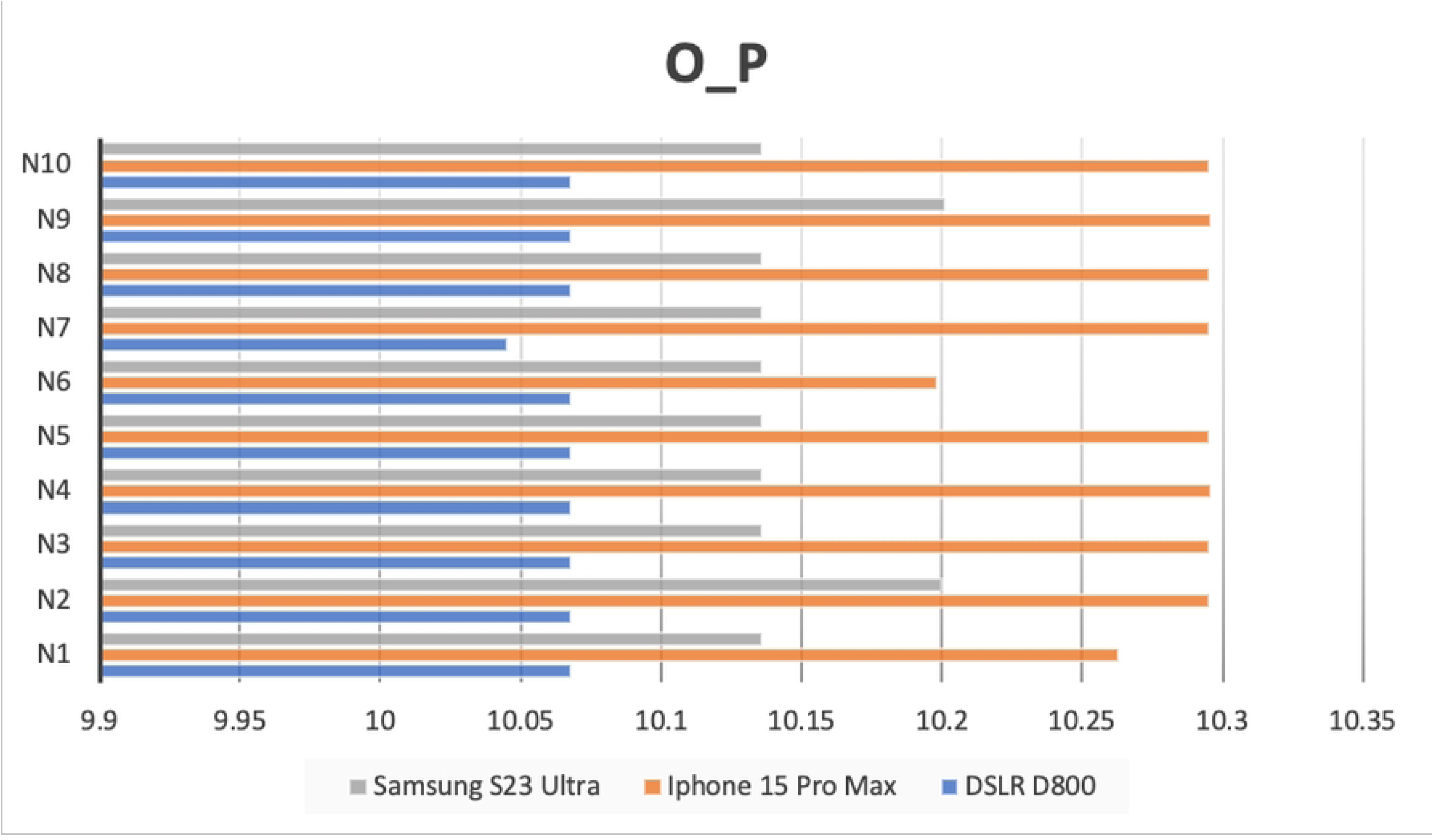
Result of O_P in occlusal view image.

**Fig 30.**
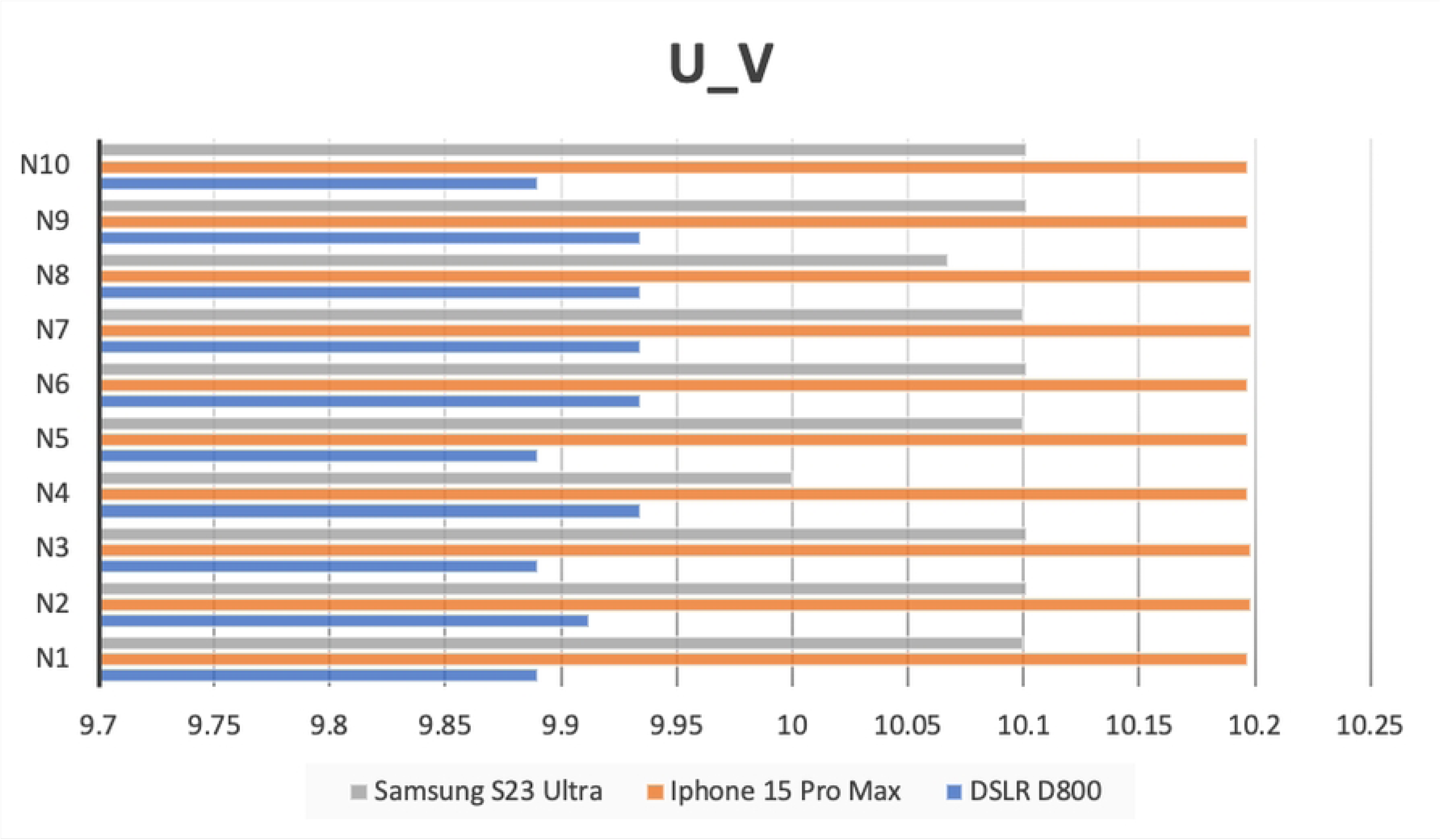
Result of U_V in occlusal view image.

**Fig 31.**
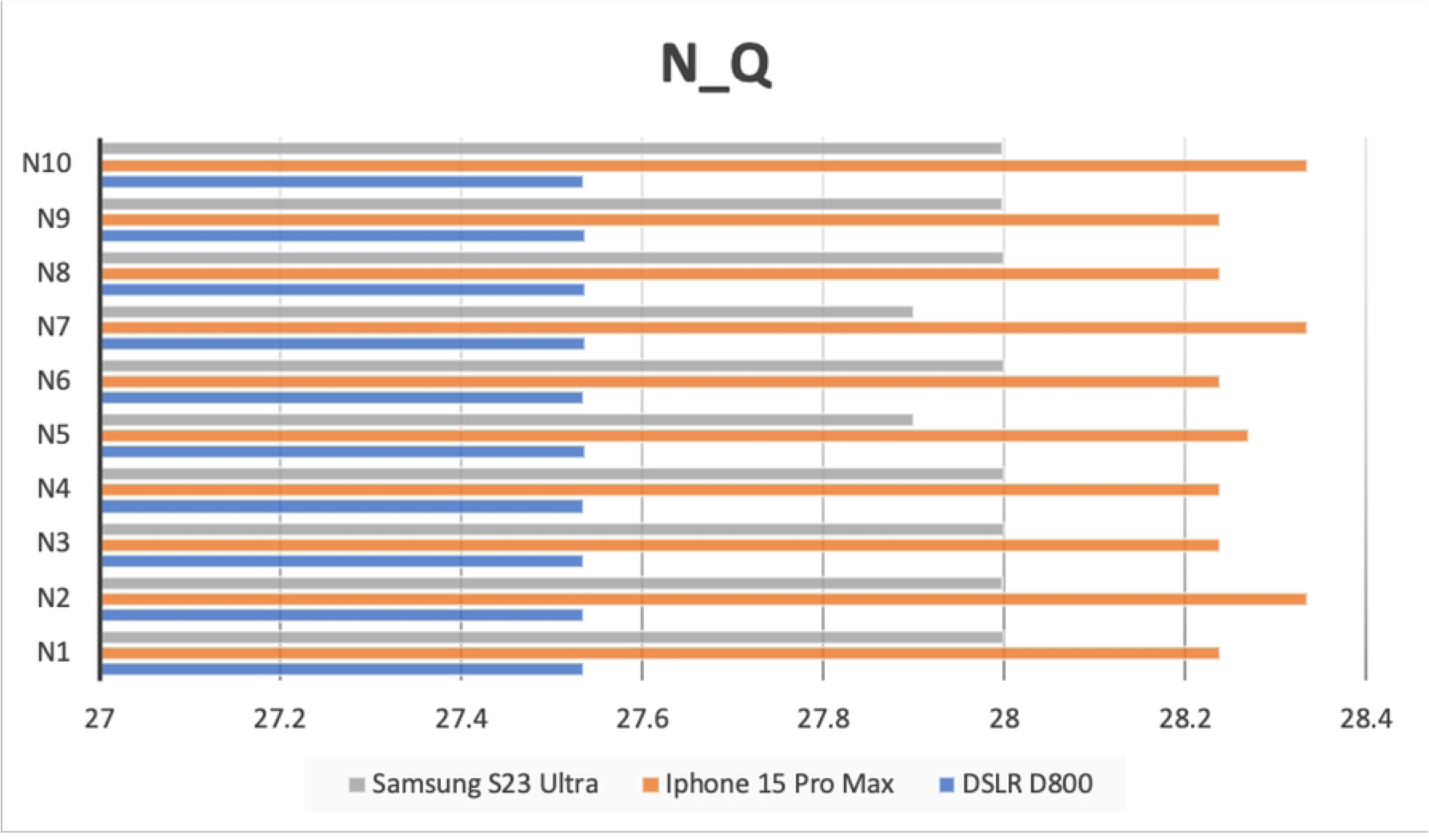
Result of N_Q in occlusal view image.

**Fig 32.**
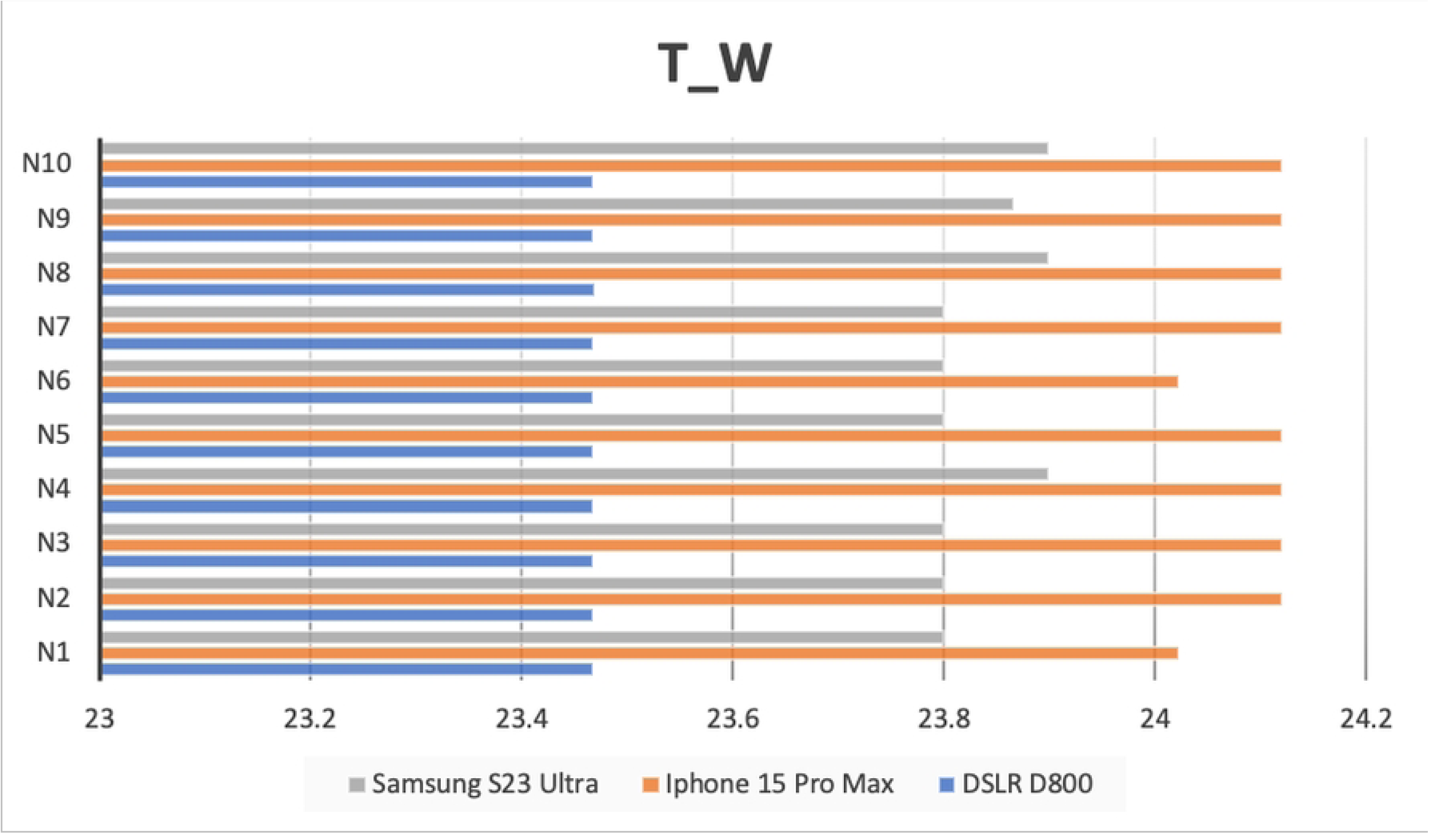
Result of T_W in occlusal view image.

**Fig 33.**
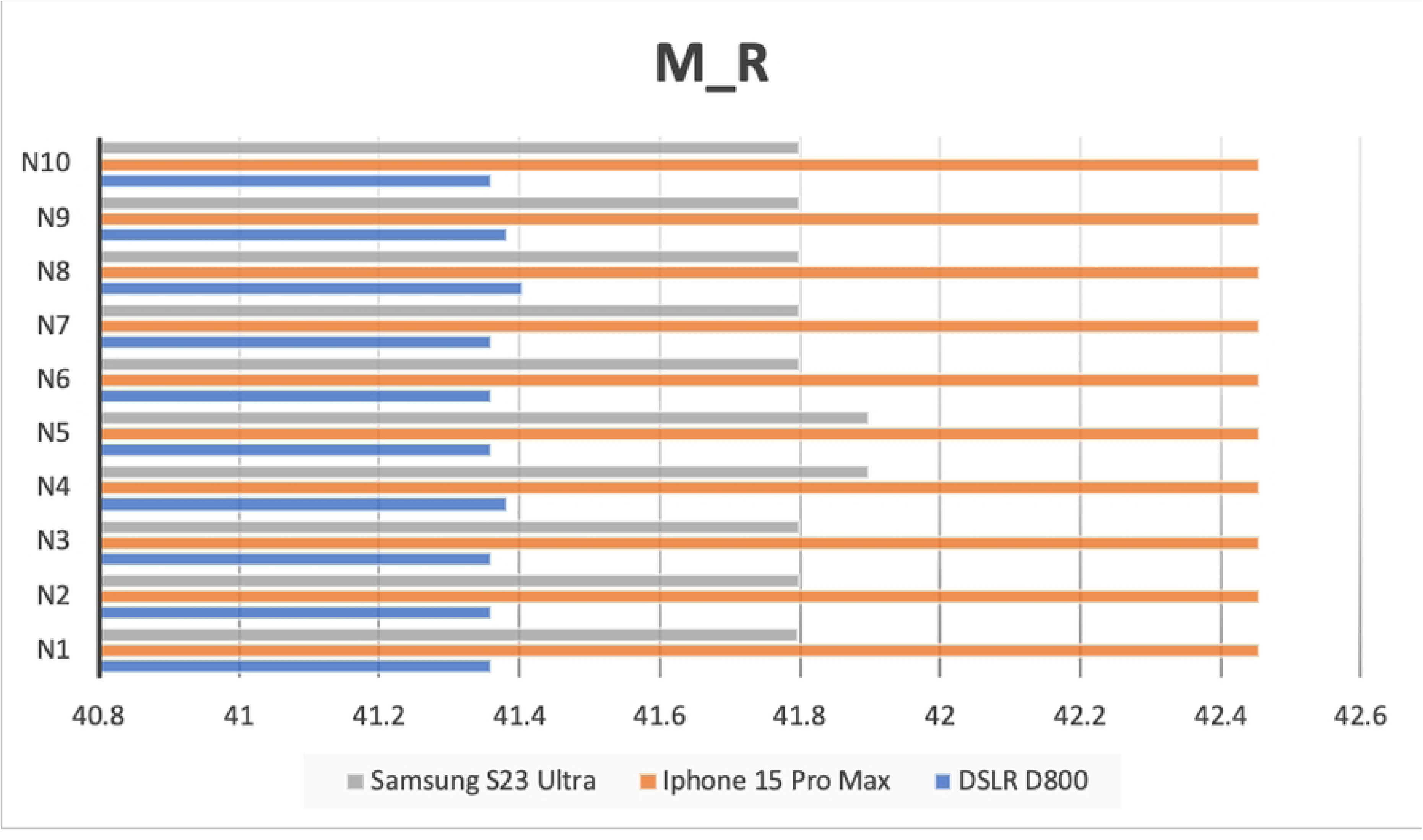
Result of M_R in occlusal view image.

**Fig 34.**
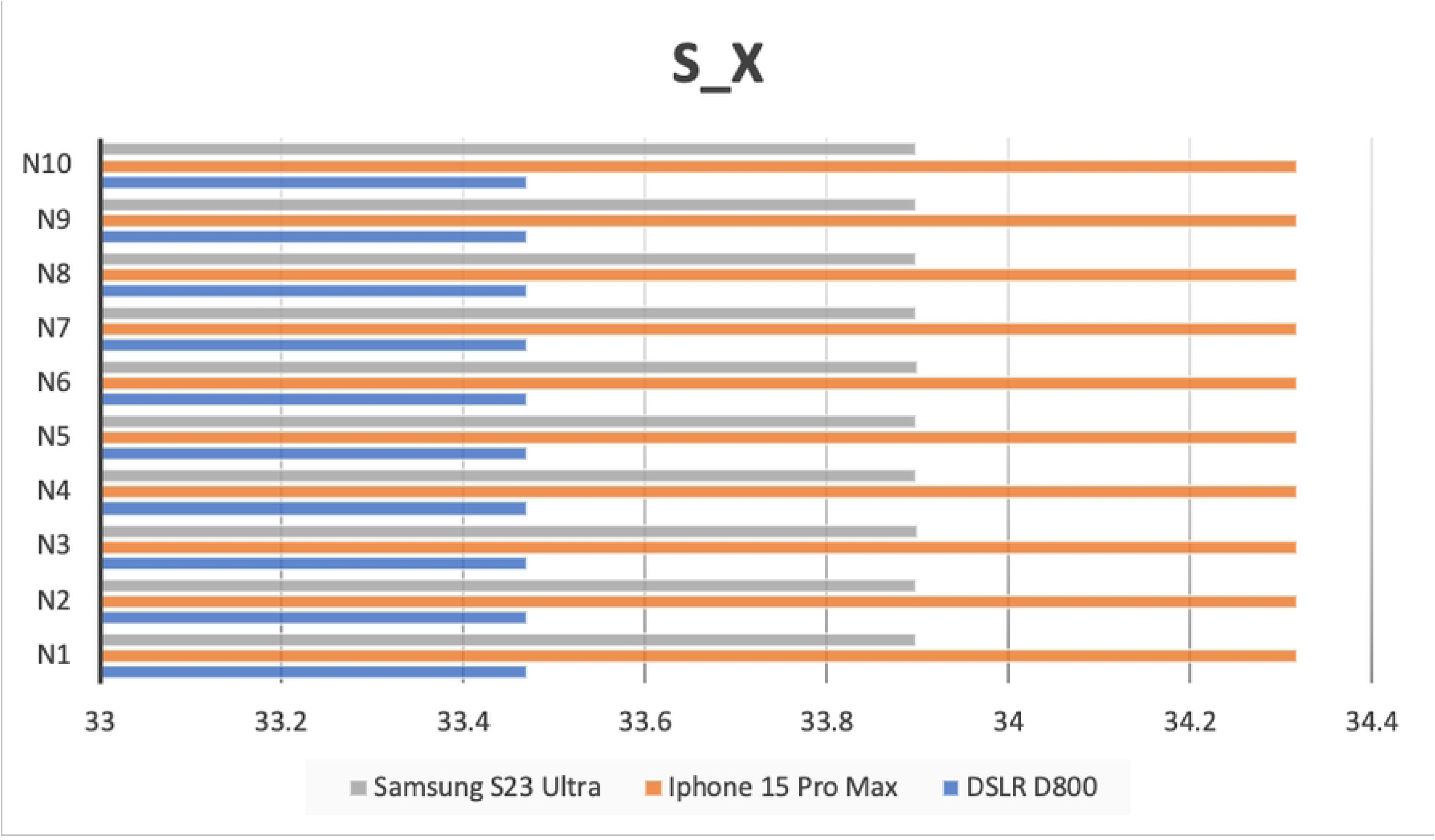
Result of S_X in occlusal view image.

## 6. DISCUSSION

This study aimed to evaluate dentists’ perceptions of intraoral images captured using a DSLR and two advanced smartphones by assessing the linear dimensional accuracy in close-up aesthetic intraoral photographs. As DSLR cameras are widely regarded as the gold standard in dental photography, this comparison provides valuable insight into the viability of smartphone cameras as practical and accessible alternatives in clinical practice.

The results of this in vitro study clearly demonstrate that the linear dimensional accuracy of images captured by some smartphones did not significantly differ from those taken with a DSLR camera, while others showed notable deviations. This highlights the variable performance among different smartphone models when used for precise dental measurements. Moussa et al. [18] previously reported no significant difference in tooth dimension accuracy between a DSLR (Canon EOS 700D with a 100-mm macro lens) and a smartphone (iPhone X), suggesting that both devices can yield reliable linear measurements when used at a minimum distance of 24 cm. In the present study, images were captured at a standardized distance of 35 cm, with a focus on evaluating JPEG files exported directly from each device, without any post-processing.

This approach was intended to assess the native image quality and dimensional accuracy provided by each camera under consistent conditions. It is important to acknowledge that the built-in automatic image optimization in smartphones may help narrow the perceptual gap in quality between DSLR and smartphone images. The findings of this study partially support those of Moussa et al. for certain smartphone models. However, the iPhone 15 Pro Max demonstrated notable discrepancies, particularly due to apparent barrel distortion affecting linear measurements.

Unlike previous studies, this research focused on close-up intraoral imaging, specifically targeting the anterior region from upper right canine to left canine. This limited field of view resulted in less peripheral space beyond the canines, potentially intensifying distortion effects. Since the measurement points were located farther from the optical center of the image, they were more susceptible to distortion, particularly the barrel distortion effect.

Despite these challenges, the Samsung Galaxy S23 Ultra demonstrated reliable performance in capturing linear dimensions, showing no statistically significant differences compared to the DSLR camera. This suggests that certain advanced smartphones may be suitable alternatives for close-up intraoral photography, particularly in clinical scenarios that benefit from increased portability, reduced setup time, and user-friendly operation.

One of the primary challenges in clinical photography is achieving image consistency, as numerous variables such as operator technique, lighting conditions, and camera settings can significantly impact the results. To minimize these variables, the present study employed a single trained operator and maintained standardized conditions for lighting, shooting distance, and angulation throughout all photographic sessions. Prior to data collection, the operator underwent a calibration process to identify the optimal settings for each camera, thereby enhancing both the consistency and reproducibility of the captured images.

DSLR cameras, such as the Nikon D800 used in this study, are commonly paired with external ring flashes to overcome the shadowing and light obstruction caused by macro lenses. The built-in flash on DSLR systems is often inadequate due to interference from the lens body. In contrast, the ring flash delivers uniform, shadow-free illumination, making it particularly suitable for high-detail intraoral photography.

Smartphone cameras, by design, offer certain advantages in this regard. Their minimal lens protrusion and close proximity between the lens and built-in flash reduce the likelihood of light obstruction, enabling unobstructed and evenly lit images. In this study, smartphone images were captured using auto mode with the telephoto lens settings to optimize detail acquisition and mimic the standardized close-up conditions used with the DSLR camera with macro lens.

Notably, the Samsung Galaxy S23 Ultra benefited from the use of the S Pen as a shutter release, allowing for stable 10 second timer shots. This reduced hand movement and yielded high quality images. The iPhone 15 Pro Max, however, produced burst shots during its 10 second timer interval, which reduced image resolution and quality. To compensate, photographs with the iPhone were taken manually at set intervals by tapping the screen.

Even with the built-in telephoto lens, smartphone cameras are limited by a reduction in pixel resolution below 12 MP, which slightly reduces image quality and increases noise due to smaller pixels—key disadvantages of smartphone cameras [34]. The evolution of smartphone camera technology, from early low-resolution CCD sensors to modern high-resolution CMOS sensors, has significantly improved their performance [32]. Enhancements in sensitivity, software optimization, and effects on natural tone, sharpness, and clarity have enabled smartphones to produce clinically useful images, supporting their potential for acceptable use in clinical settings.

Although DSLR cameras remain superior in terms of image customization and optical clarity, the gap between DSLRs and smartphones has narrowed considerably due to the rapid advancements in smartphone camera technology [6]. The use of smartphone cameras for intraoral photography has only recently begun to emerge in the literature [12, 22]. When used correctly with appropriate settings and accessories, smartphone photography can be a powerful tool for aesthetic analysis [35].

The convenience of smartphones, such as instant image review, automatic metadata recording, ease of sharing, and the ability to quickly retake images, makes them an appealing alternative in everyday dental practice [36]. Additionally, their compact size, user-friendly operation, and seamless integration into daily workflows make them highly accessible tools for documentation, patient education, and case communication. Notably, smartphones are particularly beneficial for practitioners in remote areas who need to consult with dental schools or medical centers, offering a practical solution for real-time communication and collaboration [37].

Previous research by Kathryn et al. [38] indicated that both general dentists and orthodontists showed a greater preference for intraoral photographs taken with smartphones over those captured with DSLR cameras. Similarly, another study [39] found no significant difference in image quality or characteristics between smartphone cameras (iPhone 7 Plus, Oppo R7 Plus) and a DSLR (Canon 700D), based on questionnaire responses from general dentists and orthodontists.

In dental specialties, smartphones are increasingly being utilized as preliminary screening tools in both dental and orthodontic settings [40]. However, when choosing the appropriate camera for clinical use, factors beyond photographic quality—such as accessibility, ease of use, and integration into workflow—must also be considered.

This study aimed to serve as a foundational step for future research on the application of smartphones in dental photography. However, certain limitations must be acknowledged. The study was subject to selection bias, as it involved only a single study model and focused exclusively on horizontal linear measurements. Ideally, a broader range of anatomical diversity and measurement dimensions would strengthen the generalizability of the findings.

Moreover, the inclusion of the Samsung Galaxy S23 Ultra and iPhone 15 Pro Max was intended solely as a representative examples of advanced smartphones and does not reflect the performance of all smartphone models. Therefore, the linear dimensional accuracy observed in these devices should not be interpreted as indicative of the overall capabilities of smartphone cameras in dental photography.

## 7. CONCLUSION

Within the limitations of this study, the following conclusions can be drawn. The Samsung Galaxy S23 Ultra demonstrated no statistically significant differences in linear dimensional measurements when compared to the DSLR camera, indicating its potential suitability for clinical dental photography. Conversely, the iPhone 15 Pro Max exhibited slight deviations.

Nevertheless, mobile photography is becoming an increasingly integral component of clinical dental practice. With ongoing technological advancements, further enhancements in smartphone camera systems are expected, which will improve their reliability and expanding their utility in dental applications. When used under controlled conditions and with appropriate calibration, smartphone photography represents a promising, efficient, and practical approach to intraoral imaging. It has the potential to significantly aid in treatment planning, enhance patient communication, and streamline clinical documentation. The current limitations of dental photography using smartphone cameras may be addressed and overcome through continued innovation and research.

## Data Availability

All relevant data are within the manuscript and its Supporting Information files.

## 8. ACKNOWLEDGEMENTS

The success of this thesis can be achieved by the attentive support from

Dr. Surakit Visuttiwattanakorn, and Dr. Parinya Amornsettachai. Their expertise, insightful comments, and dedicated advice throughout the process have been instrumental. Their dedication and time have been precious to me. Their support motivated me to consistently dedicate effort to my research project.

I would also like to extend my gratitude to all the respondents who participated in this study. Additionally, I would like to thank the Mahidol digital dental center for their support, both in providing personnel who offered guidance on the use of equipment and tools, as well as for supplying the necessary resources and instruments essential to this research.

## Supporting information

**S1_Table. Supporting information for Table 4 and 5.**

**S2_Table. Supporting information for Table 4 and 5**

